# Small-sample estimation of the mutational support and the distribution of mutations in the SARS-Cov-2 genome

**DOI:** 10.1101/2020.04.23.20076075

**Authors:** Vishal Rana, Eli Chien, Jianhao Peng, Olgica Milenkovic

## Abstract

The problem of estimating unknown features of viral species using a limited collection of observations is of great relevance in computational biology. We consider one such particular problem, concerned with determining the *mutational support and distribution* of the SARS-Cov-2 viral genome and its open reading frames (ORFs). The mutational support refers to the unknown number of sites that is expected to be eventually mutated in the SARS-Cov-2 genome. It may be used to assess the virulence of the virus or guide primer selection for real-time RT-PCR tests during the early stages of an outbreak. Estimating the unknown distribution of mutations in the genome of different subpopulations while accounting for the unseen may aid in discovering adaptation mechanisms used by the virus to evade the immune system. To estimate the mutational support in the small-sample regime, we use GISAID sequencing data and new state-of-the-art polynomial estimation techniques based on weighted and regularized Chebyshev approximations. For distribution estimation, we adapt the well-known Good-Turing estimator. We also perform a differential analysis of mutations and their sites across different populations. Our analysis reveals several findings: First, the mutational supports exhibit significant differences in the ORF6 and ORF7a regions (older vs younger patients), ORF1b and ORF10 regions (females vs males) and as may be expected, in almost all ORFs (for Asia versus Europe and North America). Second, despite the fact that the N region of SARS-Cov-2 has a predicted 10% mutational support, almost all observed mutations fall outside of the two regions of paired primers recommended for testing by the CDC.

**Author Summary:** We introduce the new problem of small-sample estimation of the number of mutations and the distribution of mutations in viral and bacterial genomes, and in particular, in the SARS-Cov-2 genome. The approach is of interest due to the fact that it aims to predict which regions in the genome will mutate in the future and with what frequency, given only a very limited number of complete viral sequences. This setting is usually encountered during the early stages of an outbreak when it is critical to assess the potential of the virus to gain mutations advantageous for its spreading. The results may also be used to guide the selection of genomic (primer) regions that are not subject to mutational pressure and can consequently be used as identifiers in the process of testing for the disease. They can also highlight differences in the mutation rates and locations of the SARS-Cov-2 virus affecting diverse subpopulations and therefore potentially suggest the role of certain mutations in evading the immune system. Our approach uses a new class of estimation methods that may find other applications in bioinformatics.

## Introduction

Viruses mutate for a number of reasons, due to unreliable replication of their genetic content and due to their need to evolve, adapt and evade the immune system of the host organism. The rate of mutation varies widely across various types of viruses and has been extensively studied in the past [1, 2]. It is known that RNA viruses tend to mutate faster than DNA viruses as RNA replication is much less accurate than DNA replication. Similarly, single stranded viruses are more likely to mutate than double stranded ones [3] due to structural instabilities. There is also evidence to indicate that the length of the viral genome is inversely correlated with the mutation rate, with shorter viruses mutating faster than those having longer genomes [4].

Mutational and fitness landscapes of viruses are frequently used to assess their potential to spread within diverse populations [5–7]. If the immune system of a host encounters viral proteins that it was already exposed to, its response is fast and the infected cells are efficiently eliminated. If the virus mutates at a very high rate, the host immune system may take longer to respond, giving the virus more time to replicate and spread. This phenomenon is known as *antigenic drift* [8] and fast mutating viruses pose a greater health risk as they have escape mechanisms not countered by the host [9, 10]. Some recent studies have shown that high mutation rates can also trigger a rapid innate immune response in the host; hence, they can also be a sign that the host is successfully fending of the infection and that the virus has to explore a significant number of changes in its genome to successfully compete with the immune system. On short time scales, elevated levels of mutations may even be disadvantageous to the survival of viruses [11]. Despite all these insights, it is still a challenging problem to determine the exact causes of elevated mutation rates in some viruses and their correlation to clinical patient outcomes. The first step in addressing this and other related issues is to accurately estimate the mutation rates and the distributions of the mutations.

The definition of a viral “mutation rate” varies significantly [1, 12]. What is referred to as the *genomic mutation rate* is the product of the per-nucleotide site mutation rate and the genome size, and it represents the average number of mutations each viral offspring has with respect to the parental (or ancestral) genome. RNA viruses have a per site mutation rate that lies in the range 10^−6^ − 10^−4^ [12]. The mutation rate of a virus is also often equated with the rate at which errors are made during replication of the viral genome. Nevertheless, it is clear that replication errors are not the only mechanism behind viral mutations. Other estimates are based on counting the mutations in sequenced genomes, using a reference corresponding either to Patient 0 (the first infected individual) or more frequently, to Patient 1 (the first individual that was sequenced). In the former context, the genome mutation rate for SARS-Cov-2 is estimated to be 2-3 mutations a month [13]. Defining the genomic mutation rate of a large carrier population is an even more challenging task as each host may harbor viruses with widely different mutation rates.

We define the mutational support of a virus as follows. First, we declare the viral genome of Patient 1 or some other patient as a reference and index all locations along the genome. The mutational support of a single viral genomic sequence equals the set of locations where it disagrees with the reference. The size of the mutational support hence equals the Hamming distance between the reference and the sequence under consideration. The *mutational support of a population (henceforth, mutational support)* of viral genomes equals the size of the union of the individual mutational supports. One cannot expect to be able to directly observe the mutational support of a population as not all patient’s viral genomes are sequenced and as the mutations may change in time. To estimate the mutational support, one can use a limited number of samples and count the the total number of genetic sites mutated in at least one viral genome. Counts (or maximum likelihood estimators) only offer good estimates of the actual mutational support when the number of genomic samples is significantly larger than the length of the viral genome. In other words, simple counting of mutations when only a small number of sequenced genomes are available may produce highly inaccurate estimates due to unseen mutations (caused by not having sequenced every individual and by not being able to account for all mutations). The small-sample effect is a well-known phenomena extensively studied in the machine learning community [14, 15]. Nevertheless, to the best of the authors knowledge, the problems of mutational support and mutational distribution estimation in the small-sample regime have not been addressed in the virology literature. We argue that this problem is of significant relevance as its successful solution may be used to assess the virulence of the virus, guide primer selection for real-time RT-PCR tests during the early stages of an outbreak and correlate mutational rates with elevated risks of heavy symptoms.

Our contributions are two-fold. First, we present new machine learning methods for determining the unknown support of mutations and their distributions given sequencing data from a limited number of Covid-19 patients. The methods use efficient polynomial class estimators and exhibit state-of-the art performance on synthetic datasets. The actual genomic datasets are retrieved from the Global Influenza Surveillance Aid (GISAID) repository during the early stages of the Covid-19 outbreak. In our initial analysis, we only use < 9, 000 samples, which is a significantly smaller number than the length of the SARS-Cov-2 genome which roughly equals 30, 000. The approach is based on weighted Chebyshev polynomial estimators and adapted Good-Turing distribution estimators, and its accuracy is evaluated based on larger sample set sizes retrieved on later dates. Second, the mutational supports are estimated for three different population types, namely according to geographic region (Asia, Europe, North America (NA)), gender (female/male) and age (< 55, > 55). For European samples retrieved at a later time stage, estimates for females of age < 55 versus males of age > 55 were analyzed as well. The estimates are used to predict mutational hotspots and compare the genomic loci with highest mutation frequency in different subpopulations. For the latter task, we further process the results by using the Jaccard distance as well as the symmetric Kullback-Leibler divergence. Furthermore, to determine if the mutation rates are appropriately low in genomic regions harboring primers used for real-time reverse-transcriptase polymerase chain reaction (RT-PCR) testing [16], we separately scrutinize the N ORF of SARS-Cov-2 samples.

Our analysis reveals several important biological findings. The predicted mutational supports exhibit significant differences in the ORF6 and ORF7a regions in older versus younger patients, ORF1b and ORF10 regions in females versus males. The mutational support of the ORF1b region for young females is almost twice that of old males, while old males have a significantly larger mutational supports for genes S and ORF10. Given that young females are much less likely to develop severe symptoms than old males, the identified potential high-mutation regions may be further examined to identify their potential role in the spread and severity/potency of the virus. Furthermore, it is important to observe that the variance of the support is extremely high in the ORF8 region, close to 200 times higher for patients above 55 years of age compared to patients below 55 years of age. Less surprisingly, there also exist statistically significant differences in the ORFs of Asian versus European and NA samples in the ORF1a,b and other ORFs. Second, despite the fact that we predict that the N region of SARS-Cov-2 will have a very large mutational support, almost all high-probability mutations fall outside of the two regions of paired primers recommended by the CDC for RT-PCR testing.

Note that the observed frequencies of mutations across viral genomic sites are a function of numerous and complex factors that are still poorly understood; they include population dynamics, viral-host interactions and natural selection. They also reflect the random timings at which mutations occur. As one can expect, given the lack of adequate models, our analysis and synthetic data simulations cannot completely account for the above described phenomena. Nevertheless, our methods for sequence analysis have provable guarantees for some simplified models, which is seldom the case for computational biology methods. Furthermore, mutations inherently have an underlying distribution which may change in time; to account for this issue, we address the dynamics of the mutation process by sampling genomes made available at different times and comparing the prediction results based on earlier (and smaller) time-stamped collections with those actually observed at a latter point in time.

The paper is organized as follows. In the Materials and methods Section, we describe the data acquisition process, the pre-processing tasks as well as our new small-sample support and distribution estimation algorithms. The most relevant results and the discussion of their biological relevance are presented in the Results and Discussion Section.

## Materials and methods

### Organization of the SARS-Cov-2 genome

A breakdown of the genomic structure of SARS-Cov-2 is depicted in Figure 1, and described in detail in [17] and [18]. Understanding the roles played by various ORFs of the viral genome is of importance as it allows one to put the results of the mutational support analysis into proper context: Mutational variability in certain ORFs of different host subpopulations may be indicative of different innate immune responses and evading mechanisms employed by the virus.

**Figure 1:**
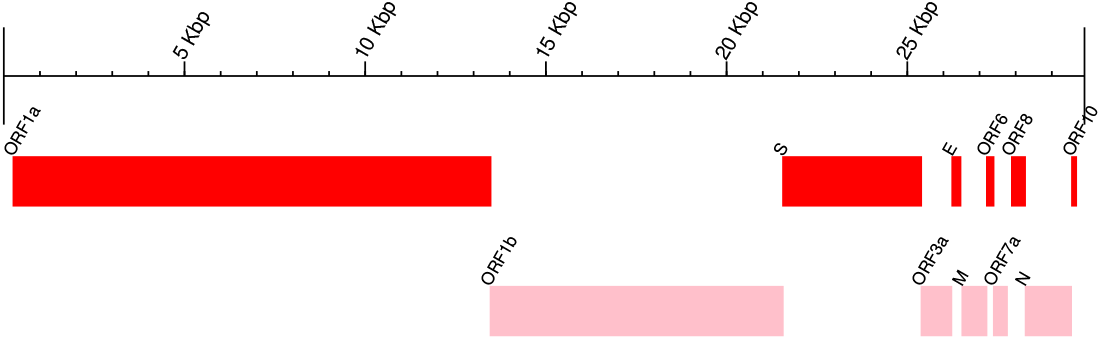
Organization of the SARS-Cov-2 genome. (Wuhan-Hu-1, GenBank MN908947)

Typically, coronaviruses have genomes including at least six open reading frames (ORFs). ORF1a and ORF1b constitute the longest component of the genomes and are responsible for encoding two polypeptides, pp1a and pp1ab, which are jointly used to create a family of nsp proteins. This family of polypeptides includes replicase-transcriptase proteins, responsible for promoting cellular mRNA degradation and blocking the translation process in host cells, thereby impairing the operation of the immune response and proofreading. The pp1a/b polypeptides are functionally combined using proteases, such as the native chymotrypsin-like protease. Viral structural proteins are encoded by the sgRNA region, and include the ORF2 or spike (S), ORF5 or membrane (M), ORF4 or envelope (E), and nucleocapsid (N) proteins, as well proteins encoded by the ORF10 sequence. ORF3a encodes a membrane protein that interacts with proteins encoded by ORFs M, S and E and is believed to play an important role in viral release and the generation of cytokine storm; on the other hand, ORF3b encodes protein that block the induction of interferons with antiviral activity. The ORF6 products are important virulence factors that enable the virus to escape detection by the immune system of the host.

For real time RT-PCR testing and detection of Covid-19, the oligonucleotide primers and probes are selected from the nucleocapsid (N) gene region (per CDC guidelines for the United States [19]), and as provided in panels produced by Integrated DNA Technologies (IDT), including two primer pairs/probe sets. As a control, additional primer/probe sets are used such as the human RNase P gene (RP) which is also included in the panel. Countries like Germany and China have adopted primers from other genomic regions, as outlined in [16]. For individual testing for Covid-19 in the United States, it is of special interest to predict mutation rates in the N region of the genome [16]. High-rate mutations in this region may cause highly undesirable false negatives in the test outcomes. ORF7a encodes for a membrane protein while ORF7b is believed to act as a viral attenuation factor and contributor in human infectivity, similarly to the protein encoded by ORF8. The ORF9b has the role to impede mitochondrial morphology and function and disable the interferon response of the host, while ORF9c appears to block important signaling pathways of the host [18].

### Data acquisition

For the proposed analyses, we used data from the GISAID EpiCoV database [20] which contains sequenced viral strains collected from patients across the world. We downloaded the data at three time points, starting from 04-03-2020, continuing on 04-10-2020 and finishing on 04-14-2020. We then revisited the repository on 10-20-2020 to further evaluate the quality of our predictions regarding the mutational supports. At that point of time, 9, 271 samples from Asia and more than 30, 000 samples from NA and 85, 000 samples from Europe were available.

For samples made available in April as well as in September 2020, we filtered the datasets only to include nearly-complete samples i.e., those of length > 29, 000 nts, resulting in a number of samples summarized in Table 1. We also downloaded the associated metadata used for patient subtyping. Note that we used results obtained early in the monitoring process in order to evaluate our small-sample estimation schemes. Table 1 provides the number of samples available within different categories for each of the three time points.

**Table 1:**
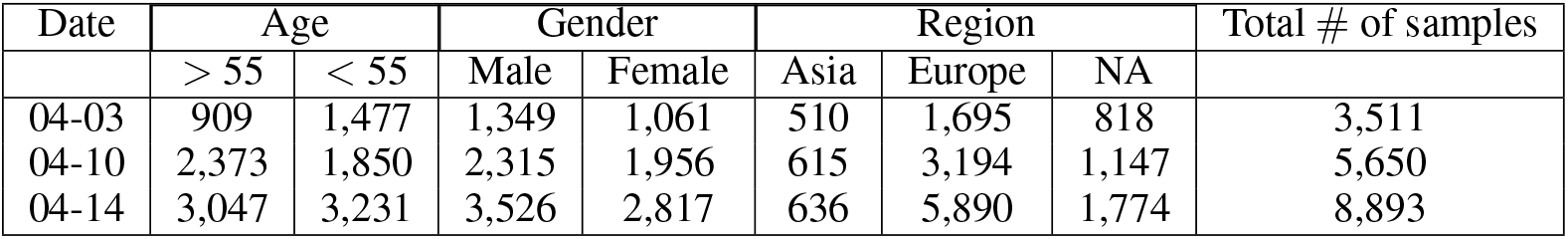
Number of samples available for different phenotype classes and data retrieved on three different dates, 04-03-2020, 04-10-2020 and 04-14-2020.

As the first step in our analysis, we used the sequence alignment software MUSCLE [21] to perform pairwise alignment of all the samples with the SARS-Cov-2 sequence of Patient 1, published under the name Wuhan-Hu-1, admitted to the Central Hospital of Wuhan on December 26, 2019 (GenBank accession number MN909847). Furthermore, we also performed alignment with respect to Patient 1 of two additional continents, Europe and NA. The latter alignment was performed to better determine how the mutational support and mutational distribution depends on a particular geographic context.

For each aligned pair of samples, we generated a “mutation profile”, a list containing the positions in the reference genome in which the patient aligned to the reference has a substitution mutation. We did not perform multiple sequence alignment in order to assess the mutation landscape as we need to analyze each patient data separately (each patient and her/his mutations are treated as one sample in the estimation procedure). The mutational profile lists are subsequently aggregated over all the patient samples, resulting in a histogram of mutations across all positions in the viral reference genome. The aggregate profiles are further partitioned according to the 11 genes they are located in on the viral genome depicted in Figure 1. The total count of mutations for each location in each gene is used as a sufficient statistics for estimating the mutational support and the distribution of the mutations in each of the 11 genes. The analytic pipeline used is depicted in Figure 2.

**Figure 2:**
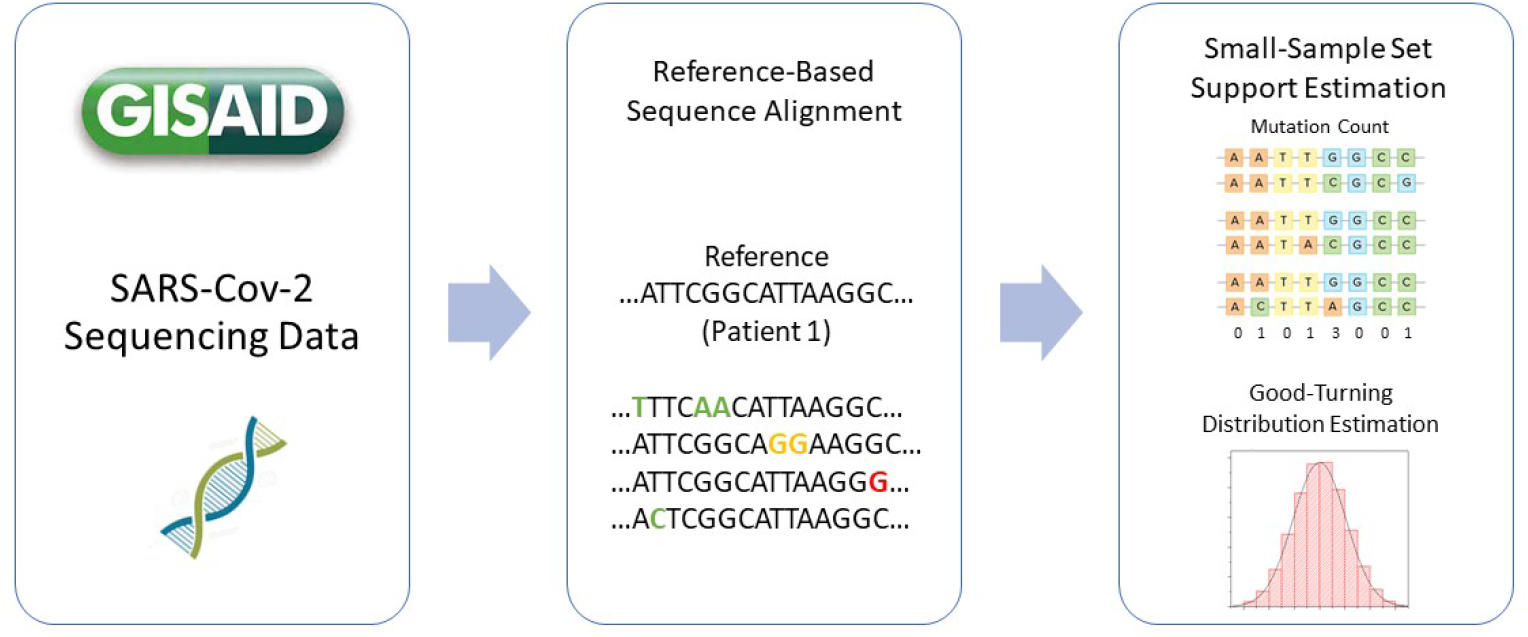
The data analysis flowchart: Viral sequencing data is retrieved from the GISAID repository and then aligned against the genome of Patient 1 or regional Patient 1 in a pairwise fashion. The substitutions at different genomic locations for all analyzed pairs of samples are counted and used as sufficient statistics for the estimation procedures.

To adjust for alignment artifacts introduced by sequencing errors, dropouts and alignment gaps, we removed all gaps encountered in the prefixes and suffixes and sufficiently long gaps (> 10 nts) within the actual alignments. Most gaps are encountered at the 5’UTR and 3’UTR regions of the genome, as may be expected from outputs of global alignment algorithms.

As there exists a large body of evidence of stratified susceptibility and severity of symptoms across different racial, age and gender groups [22, 23], we performed four different types of mutational support and distribution analyses. In the first set of tests, we split the patient mutation histograms based on gender (male/female), based on age (under 55/over 55) and based on the geographic location (Asia/ NA/ Europe). The age threshold was set by taking into consideration available sample sizes needed for the analysis and the age profile of patients available on GISAID; the threshold also reflects different risk groups for the development of severe symptoms. In addition, we performed the same analysis for a combination of patient features for settings with sufficiently many samples available early in the pandemic, such as males above 55 years of age/females below 55 years of age, from Europe. Note that in all the described cases, “geographic location” refers to the region of infection of the patient and not the region where he/she was tested and the sample was sequenced.

Since the number of samples per population type may vary significantly, we performed two tests. In one test we used all samples available, while in another we adjusted for difference in sizes of the sets by subsampling the larger of the two classes to make the sample sets of equal sizes. The number of samples available for various patient subgroups is listed in Table 1. For data obtained on 04-03-2020, we used all the samples available for all the classes, without balancing the class sizes. For data retrieved on 04-10-2020 and 04-14-2020, we balanced the classes by subsampling from the larger of the two classes for both age- and gender-based subtypes. For different geographical regions, on 04-10-2020, we used all 615 samples from Asia and subsampled Europe and NA to 1000 samples each. Similarly, we used all 636 samples from Asia and subsampled Europe and NA to 1, 774 samples each, for data retrieved on 04-14-2020. It is important to point out that by performing the experiments with different sample set sizes one can compare the quality of the estimates obtained using samples from the early stages of epidemics and those obtained from later stages when more information about individual viral sequences is available. Furthermore, the new machine learning methods outlined in the section to follow apply to any other viral or bacterial dataset collection with the obviously required modifications to account for the genetic profile of the microorganisms.

### New small-sample support estimators

We focus on the *polynomial approximation approach* put forward in [24], and significantly improve on it in practice by introducing new weighted Chebyshev polynomial optimization techniques largely unknown in the machine learning and computational biology community [25]. The weighted approximation method can be seamlessly combined with regularization techniques that use the variance of the estimator in a way that complements features used in Maximum Likelihood (ML) estimation [26]; and with Semi-Infinite Programming (SIP) solvers that produce the parameters of the estimator. The SIP methods can be solved consistently and highly efficiently through discretization resulting in a small Linear Program (LP) of size *decreasing with the number of samples*. Interestingly, despite the fact that our estimators are constructed using an LP as is the case for the best performing ML-based approach [27], the ML-LP formulation has a number of variables and constraints that actually *increases* with the number of samples; this difference makes our estimator significantly more efficient as is needed for large scale estimation processes like the ones described in this work, in addition to improving their performance.

Next, we provide a detailed description of our polynomial estimation method. Recall that the support of a discrete probability distribution is defined as the number of symbols with positive probability of occurrence. We define the mutational support of a virus as the total number of genomic sites mutated in any viral genome in any individual (observed or unobserved due to limited testing), compared to a reference genome. As already pointed out, in our case the reference is the genome of Patient 1, the first sequenced SARS-Cov-2 genome or the genome of regional Patient 1.

The most commonly used techniques for support and distribution estimation are ML methods which directly use the empirical counts of the symbols to determine the support or probabilities of interest. It is well known that ML approaches perform poorly for large alphabet sizes (supports) when only a small number of samples from the distribution is available. In this case, they fail to take account for samples that have never been observed due to limited sampling. To see why this is the case, assume that we observe 10 samples from a distribution supported on {1, …, 100}. Clearly, with only 10 samples available, our best possible guess for the support size will be the number of distinct symbols observed which is a number ≤ 10 and far from the correct value 100.

The problem of estimating the support of an unknown probability distribution or estimating the distribution itself in the context of small-sample sets has a long history. The first line of work in this area is attributed to Laplace, as described in [28], who introduced a class of smoothed distribution estimators termed add 1 (or more generally, add constant *c* estimators). These estimators adjust the counts of observed symbols in order to account for the unseen symbols.

Let *P* = (*p*_1_, *p*_2_, …) be a discrete distribution over some finite alphabet and let **x**^*n*^ be a vector of i.i.d samples drawn according to the distribution *P*. The problem of interest is to estimate the support size, defined as 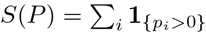. We use *S* instead of *S*(*P*) to avoid notational clutter. An important assumption used in our estimation methods is that the minimum non-zero probability of the distribution *P* is greater than 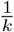, for some *k* ∈ ℝ^+^, i.e., inf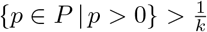. We let *D*_*k*_ denote the space of all probability distributions satisfying inf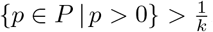. A sufficient statistics for **x**^*n*^ is the empirical distribution (i.e., histogram) *n* = (*n*_*1*_, *n*_*2*_, …), where 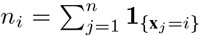 and **1**_*A*_ stands for the indicator function of the event *A*.

To determine the quality of an estimator, we use the most-frequently studied risk model, the minmax risk under normalized squared loss, defined as

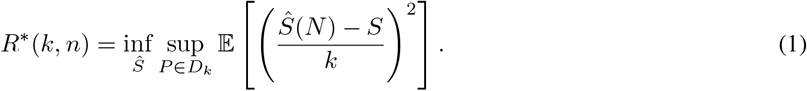

We seek a support estimator *Ŝ* that minimizes

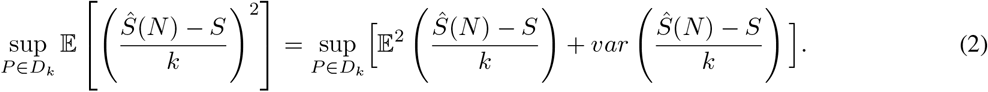

The first term within the supremum captures the expected bias of the estimator *Ŝ*. The second term represents the variance of the estimator *Ŝ*. A”good” estimator should jointly balance out the worst-case contributions of the bias and variance (note that for the case that only the bias is considered directly, and the variance accommodated for by modifying the bias-optimized solution, the underlying estimator was analyzed in [24]).

To introduce our method, we first describe the class of *polynomial estimators*. Given a positive integer parameter *L*, we say that an estimator *Ŝ* is a polynomial class estimator with a threshold parameter *L* (i.e., a *Poly*(*L*) estimator) if it takes the form *Ŝ* = ∑ _*i*_ *g*_*L*_(*n*_*i*_), where *g*_*L*_ is defined as

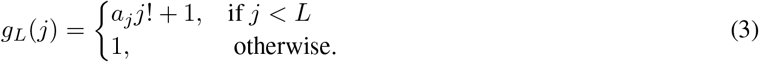

The coefficients *a* satisfy *a*_*j*_ ∈ ℝ, and *a*_0_ = −1, (since this choice ensures that *g*_*L*_(0) = 0) and have to be optimized in order to minimize the risk. One can associate an estimator *Ŝ* with its corresponding coefficients **a**, i.e.,

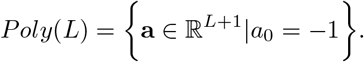

The authors of [24] proposed using a special form of polynomial estimators in which the coefficients *a*_*j*_ correspond to scaled evaluations of a Chebyshev polynomial of order *L*. The Chebyshev polynomial of the first kind of degree *L* is defined as

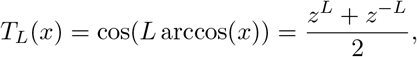

where *z* is the solution of the quadratic equation *z* + *z*^−1^ = 2*x*. The polynomial *T*_*L*_ is bounded in the interval [−1, 1] and may be scaled and shifted to lie in an arbitrary interval [*l, r*] based on

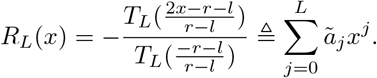

Clearly, *R*_*L*_(0) = −1 and *ã*_0_ = −1.

The Chebyshev polynomial estimator is an estimator for which

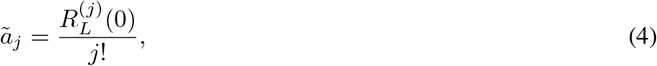

and it takes the form 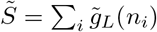, where

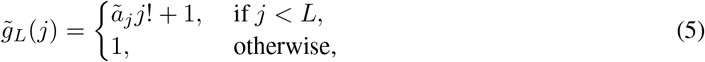

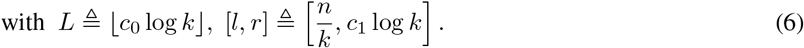

The choice values of the constants *c*_0_ and *c*_1_ are *c*_0_ = 0.558 and *c*_1_ = 0.5 and they are obtained based on an analysis of the bias and variance of the estimator.

The estimator 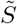 above is order-optimal *in the exponent* under the unbiased risk. Thus, the estimator can be improved by selecting coefficients of *Poly*(*L*) that jointly optimize the bias and variance term in the risk. We show how to accomplish this task by rewriting the original minmax problem as a regularized exponentially weighted Chebyshev approximation problem [25].

In order to jointly optimize the bias and variance term in the squared loss, we start by directly analyzing 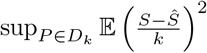. Classical Poissonization arguments lead to

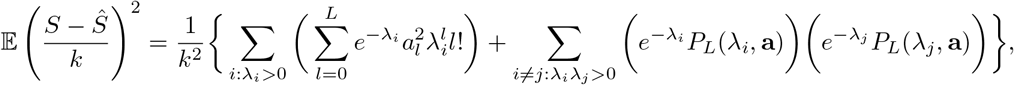

Where 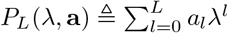. Taking the supremum over *D*_*k*_ we can bound the risk as

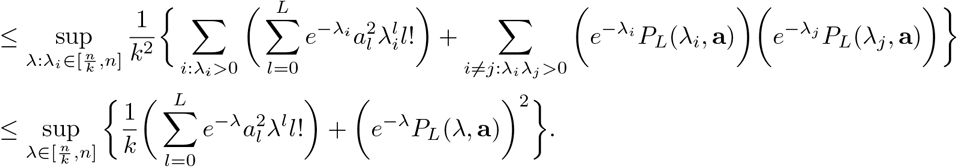

In the above inequality, we used the Cauchy-Bunyakovsky-Schwarz inequality, the fact that *S* ≤ *k* and 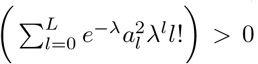, for all *λ >* 0. Hence, the optimization problem for the coefficients of the polynomial estimator at hand reads as

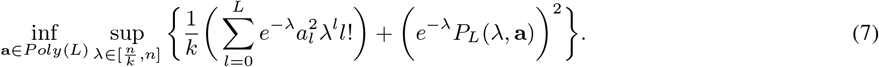

Problem (7) represents an instance of a *regularized weighted Chebyshev approximation problem*. If we ignore the first term in (7), the optimization problem becomes

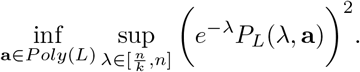

The term *e*^−*λ*^*P*_*L*_(*λ*, **a**) corresponds to the bias of the estimator. It is straightforward to see that the optimal choice of **a** for the above problem is a solution to

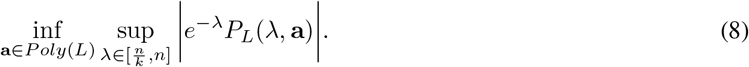

The first term 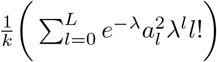,which corresponds to the variance, may be written as

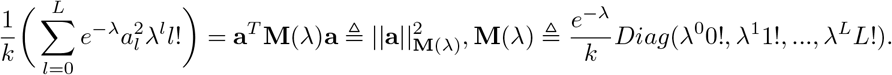

Clearly, | |.| | _**M**(*λ*)_ is a valid norm, and consequently, the first term in (7) can be viewed as a regularizer.

Simple algebra reveals that

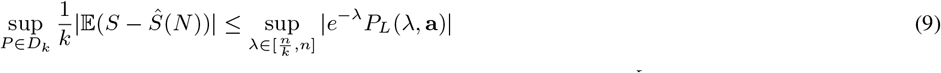

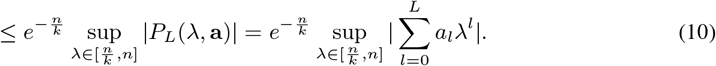

where (9) is equivalent to (8), while (10) resembles the problem studied in [24], except for a different optimization interval used within the supremum (the authors of [24] choose a shorter interval in order to decrease the contribution of the variance to the loss). Hence, optimizing (9) should produce an estimator with smaller bias as the exponential weight is inherent to the formulation. The modified bound in (10) is minimized with respect to the coefficients **a**, using the minmax property of Chebyshev polynomials [29, 30], resulting in **ã**.

To solve (7), we more closely examine some results known about weighted Chebyshev approximations [30] and semi-infinite programs. Solving for the problem directly is difficult, so we instead resort to numerically solving the epigraph formulation of problem (7) and proving that the numerical solution is asymptotically consistent.

The epigraph formulation of (7) is of the form ([31], Chapter 6.1)

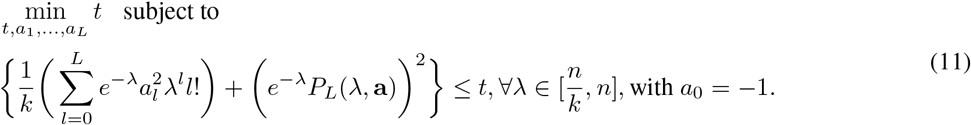

Note that (11) is a semi-infinite programming problem. There are many algorithms that can be used to numerically solve (11), such as the discretization method, and the central cutting plane, KKT reduction and SQP reduction methods [32, 33]. For simplicity, we focus on the discretization method. For this purpose, we first form a grid of the interval 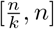 involving *s* points, denoted by Grid 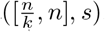. Problem (11) may consequently be viewed as an LP with infinitely many quadratic constraints, which is not solvable. Hence, instead of addressing (11), we focus on solving the relaxed problem

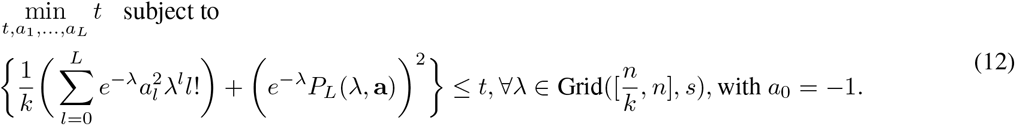

The solution of the relaxed problem is asymptotically consistent with the solution of the original problem (i.e., as *s* goes to infinity, the optimal values of the objectives of the original and relaxed problem are equal). Problem (12) is an LP with a finite number of quadratic constraints that may be solved using standard optimization tools. Unfortunately, the number of constraints scales with the length of the grid interval, which in the case of interest is linear in *n*. This is an undesired feature of the approach, but it may be mitigated through the following theorem which demonstrates that an optimal solution of the problem may be found over an interval of length proportional to the significantly smaller value log *k*, where 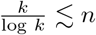 is the fundamental bound for support estimation. We relegate the proof to the Supplement.

*Theorem*. For any **a** ∈ *Poly*(*L*) and *L* = ⌊*c*_0_ log *k*⌋, and *c*_0_ = 0.558, let

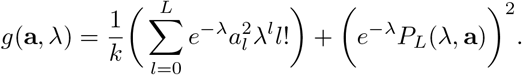

Then, we have

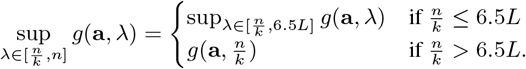

#### Remark

In weighted approximation theory [25], the problem of bounding the interval over which the supremum is achieved is a topic of significant interest, with many important available results. For example, if we ignore the regularization term, we can directly use the Mhaskar-Saff theorem to reduce the length of the interval in the supremum to 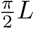. Our Theorem shows that even when a regularization term is present, we can still restrict the length of the interval to 6.5*L*. Our proof differs from that of the more general Mhaskar-Saff theorem, since we exploit the specific structure of the problem.

The optimization problem we need to solve to determine our estimator therefore reads as

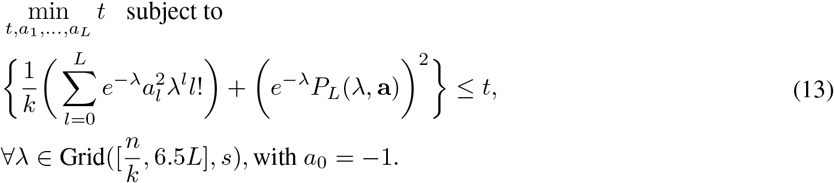

Since *L* = ⌊*c*_0_ log *k*⌋, the length of the optimization interval in (13) is proportional to log *k*.

It seems intuitive to assume that as *s* grows, the solution of the relaxed semi-infinite program approaches the optimal solution of the original problem (11). This intuition can be rigorously justified for the case of objective functions and constraints that are “well-behaved”, as defined in [34] and [35]. The first line of work describes the conditions needed for convergence, while the second establishes the convergence rate given that the discretized solver converges. We use these results in conjunction with a number of properties of our objective SIP to establish the claim in the following theorem. The proof is delegated to the Supplement.

#### Theorem

Let *s* be the number of uniformly placed grid points on the interval (13), and let 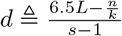 be the length of the discretization interval. As *d* → 0, the optimal objective value *t*_*d*_ of the discretized SIP (13) converges to the optimal objective value of the original SIP *t*^⋆^. Moreover, the optimal solution is unique **a**^⋆^. The convergence rate of *t*_*d*_ to *t*^⋆^ equals *O*(*d*^2^). If the optimal solution of the SIP is a strict minimum of order one (i.e., if *t* − *t*^⋆^≥*C* ‖**a − a**^⋆^‖ for some constant *C >* 0 and for all feasible neighborhoods of **a**^⋆^), then the solution of the discretized SIP also converges to an optimal solution with rate *O*(*d*^2^).

In summary, for given parameters *k* and *n*, and sample count histograms *N* we obtain the optimal coefficients of our polynomial estimators by solving the small LP program described above. An example of our polynomial estimator (henceforth termed Regularized Weighted Chebyshev (RWC) estimator) and its scaled coefficients *g*_*L*_ is shown in Figure 3, along with a corresponding example of a Chebyshev estimator (termed the Wu-Yang (WY) estimator). It is easily observed that the coefficients of the two estimators exhibit very different behaviors: Unlike the Chebyshev case, for which the coefficients have to alternate in sign, our estimators are not constrained to obey this pattern.

**Figure 3:**
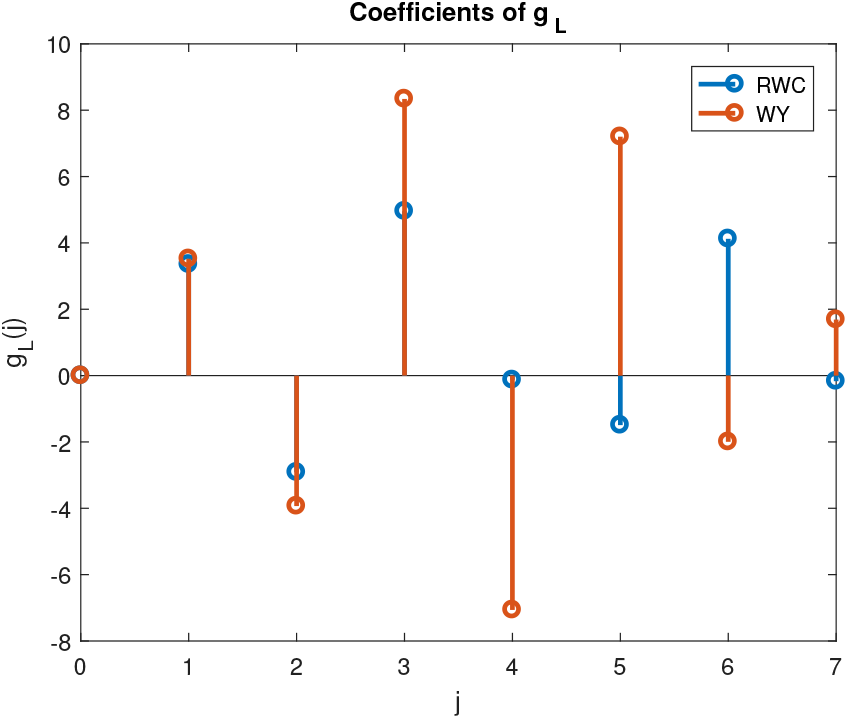
The function *g*_*L*_ for RWC and WY estimator. The parameter setting used for the illustration is *n* = *k* = 10^6^ and *c*_0_ = 0.558.

### Remark

It is important to point out that the RWC estimators are “additive”: They operate on each symbol separately and the contributions of symbols are linearly combined to obtain the overall support estimate.

We conclude by observing that our RWC estimator can be further (heuristically) improved in practice by optimizing it with respect to a minmax risk that involves a different scaling factor in the denominator. This estimator, termed the RWC-S estimator (to indicate that the scaling is performed using the result of a naive Support estimator) is described in more details in the Supplement.

### Small-sample distribution estimation

By far the most frequently used method for distribution estimation in the small-sample regime is the Good-Turing estimator [14], which tries to account for the unseen by adjusting the counts (histograms) of the actually observed symbols. In a slightly modified form the method may be described as follows. For a sequence **x**^*n*^ of length *n* over an unknown finite alphabet, we once again let *n*_*i*_ denote the number of times a symbol *i* appears in **x**^*n*^. Furthermore, we let *ϕ*_*t*_ stand for the count of counts, i.e., the number of symbols that appear *t* times in **x**^*n*^. The estimator proposed in [15] combines the Good-Turing and ML estimators, the latter being used for the frequently observed symbols. For symbols that appear *t* times, if *φ*_*t*+1_ *>* Ω(*t*), then the Good-Turing estimate is used to determine the underlying total probability mass, otherwise, the ML estimator is used instead. More precisely, for a symbol appearing *t* times, if *φ*_*t*+1_ *> t* we use the Good-Turing estimator, otherwise we use the ML estimator. If *n*_*i*_ = *t*, the estimated probability of the symbol *i* is computed according to

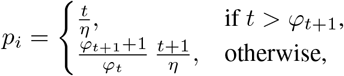

where *η* is a normalization term that ensures that the obtained values are probability masses. The term *φ*_*t*+1_ used in the Good-Turing estimator is replaced by *φ*_*t*+1_ + 1 so that every symbol has a nonzero probability.

The modified Good-Turing estimator is used instead of the classical Good-Turing estimator as the latter is known to poorly estimate the probabilities of high frequency symbols. Modifications of the Good-Turing estimator that take sampling artifacts/errors into account are also available, and implemented as described in [36, 37].

### The performance of RWC estimators on synthetic data

Consider a finite alphabet 𝒮= {1, …, *S*}. Assume that the probability of symbol *i* ∈ 𝒮 equals *p*_*i*_ and that you can randomly sample symbols from the alphabet with replacement and record the distribution histogram of *N* observed symbols. The question of interest is how accurately can one estimate *S* based on *N* samples and the parameter *k* dictating the smallest nonzero probability of the distribution.

For simplicity, assume that the alphabet is 𝒮 = {1, 2, …, 10} and that

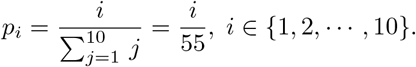

Clearly, *S* = 10 and *k* = 55. For the RWC and RWC-S estimators, we choose *L* = ⌊0.558 log *k*⌋ = 2. Now assume we draw *n* = 6 samples from the alphabet according to the specified distribution. In this case, the values of *g*_*L*_ for the RWC-S estimator are given in Table 2.

**Table 2:** The *g*_*L*_ values corresponding to the RWC-S estimator for different distinct symbol counts: Note that *Ŝ*_*c*_ denotes the naive estimator (i.e., the estimator equal to the count of different symbols).

| | $\hat{S}_c = 1$ | $\hat{S}_c = 2$ | $\hat{S}_c = 3$ | $\hat{S}_c = 4$ | $\hat{S}_c = 5$ | $\hat{S}_c = 6$ |
| --- | --- | --- | --- | --- | --- | --- |
| $g_L(0)$ | 0 | 0 | 0 | 0 | 0 | 0 |
| $g_L(1)$ | 1.8128 | 2.4819 | 2.9427 | 3.2422 | 3.4367 | 3.5758 |
| $g_L(2)$ | 1.8128 | 2.3967 | 1.7205 | 1.0699 | 0.5556 | 0.1663 |
| $g_L(j), \forall j \geq 3$ | 1 | 1 | 1 | 1 | 1 | 1 |

Consider all possible histograms of *n* = 6 symbols in this setting, summarized in Table 3. We can clearly see that except for the case *N* = [1, 1, 1, 1, 1, 1], our estimator provides a significantly better support estimation result. Note that the histogram *N* = [1, 1, 1, 1, 1, 1] arises only with very small probability (9%), and this probability significantly decreases as *n, S, k* increase. Nevertheless, even in this case, the risk (mean-square error normalized by *S*^2^) of our RWC-S estimator equals 0.2186 while that of the naive estimator equals 0.319.

**Table 3:** The estimated supports produced by the RWC-S and naive estimators for all possible histogram inputs. The probability of each histogram is computed via a Monte Carlo method with 10^6^ independent trials. Bold numbers indicate the best estimation result compared to the ground truth.

| Histogram $N$ | [1,1,2,2] | [1,1,1,1,2] | [1,1,1,3] | [1,2,3] | [1,5] | [1,1,4] |
| --- | --- | --- | --- | --- | --- | --- |
| RWC-S | **8.6242** | **14.3034** | **10.7266** | **5.6632** | **3.4819** | **6.8854** |
| Naive | 4 | 5 | 4 | 3 | 2 | 3 |
| Probability | 0.2633 | 0.3792 | 0.1374 | 0.0801 | 0.0021 | 0.0253 |
| Histogram $N$ | [2,2,2] | [3,3] | [2,4] | [6] | [1,1,1,1,1,1] | |
| RWC-S | **5.1615** | **2** | **2.7205** | **1** | 21.4548 |  |
| Naive | 3 | **2** | 2 | **1** | **6** |  |
| Probability | 0.0171 | 0.0025 | 0.0043 | 0.0001 | 0.0886 |  |

We also tested the performance of the estimators on significantly larger sets of synthetic data for which the ground truth distributions and their supports are known. In particular, we compared the RWC method with the Good-Turing (GT) estimator, the WY estimator of [24], the PJW estimator described in [38] and the HOSW estimator of [39]. We did not compare our method with the estimators introduced in [27, 40] due to their high computational complexity [39].

We considered six different distributions: The uniform distribution with 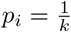, the Zipf distributions with *p*_*i*_ ∝ *i*^−*α*^, and *α* equal to 1.5, 1, 0.5 or 0.25, and the Benford distribution with *p*_*i*_ ∝ log(*i* + 1) − log(*i*). We choose the support sizes for the Zipf and Benford distribution so that the minimum non-zero probability mass is roughly 10^−6^. We run the estimator 100 times to calculate the risk. For both approximation-based estimators, we fix *c*_0_ to be 0.558. With our proposed method, we solve (13) on a grid with *s* = 1000 points on the proposed interval 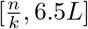. For the estimator described in [24], we set *c*_1_ = 0.5 according to the recommendation made in the cited paper. The GT method used for comparison first estimates the total probability of seen symbols (e.g., sample coverage) according to 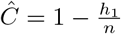, and then estimates the support size according to 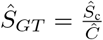; here, *Ŝ* stands for the (naive) counting estimator. Note that *h*_1_ equals the number of different alphabet symbols observed only once in the *n* samples.

Figure 4(a) shows that the RWC estimator has a significantly better worst-case performance compared to all other methods when tested on the above described collection of distributions, provided that *n* ≥ 0.2*k*. Also, both RWC and WY estimators have significantly better error exponents compared to the GT, PJW and HOSW estimators. The GT and PJW estimators perform better than RWC if 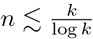, which confirms the results of our theoretical analysis as well.

**Figure 4:**
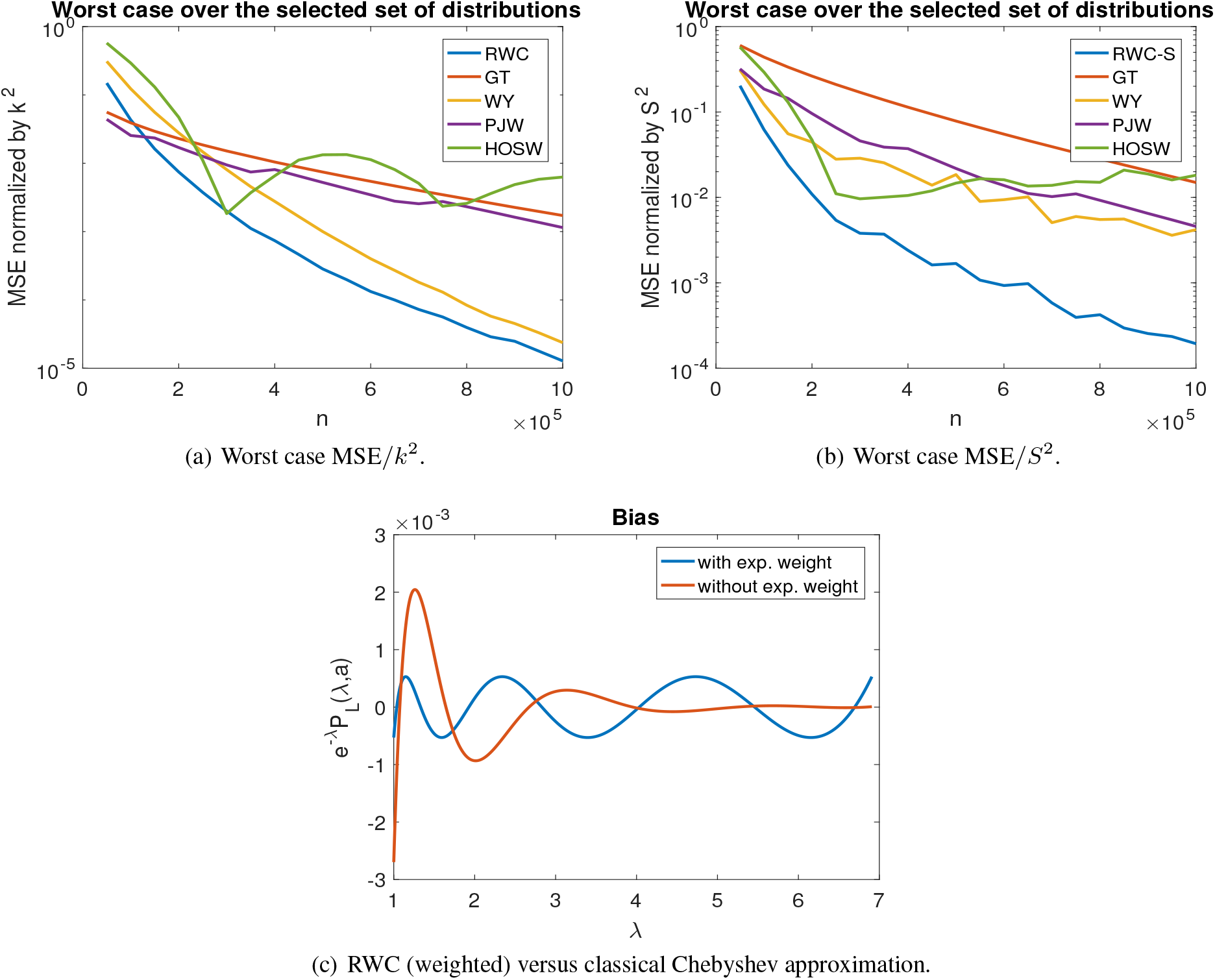
Comparison of worst case risks for different ground truth distributions and estimators. In our simulations we set *n* = *k* = 10^6^ and *c*_0_ = 0.558.

In the second set of experiments, we change the normalization from (1*/k*)^2^ to (1*/S*)^2^ as was also done in [39]. The RWC-S estimator minimizes an upper bound on the worst-case risk 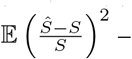 – as already pointed out, a detailed description of this algorithm and an intuitive explanation of why it outperforms the RWC method is provided in the Supplement. Figures 4(b) illustrate that our RWC-S estimator significantly outperforms all other estimators with respect to the worst-case risk normalized by *S*^2^. Moreover, the RWC-S estimator outperforms all known estimators on almost all tested distributions. As illustrated in Figure 4(c) we see that a classical Chebyshev approximation introduces a larger bias than our RWC method whenever the underlying distribution is close to uniform (i.e., when 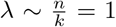). This phenomenon persists even when regularizations is taken into account.

Another common approach to testing support estimators on real data is to estimate the number of distinct words in selected books [24, 27]. Books are chosen as ground truth test cases as the words in a text are not independent and identically distributed (iid) and hence provide a means to test the performance of estimators optimized for iid settings. The performance of our approach and those of prior works on Hamlet and Macbeth can be found in the Supplement. In the experiments, we randomly sampled words in the text with replacement and used the obtained counts to estimate the number of distinct words. For simplicity, we set *k* to the total number of words. For example, as the total number of words in Hamlet equals 30, 364, we set *k* = 30, 364. Once again, our method significantly outperforms all other competitive techniques both in terms of convergence rate and the accuracy of the estimated support for all experiments.

The details about data acquisition pipeline, alignment software and implementations of the RWC and RWC-S algorithms may be found at the following GitHub repository: https://github.com/rana95vishal/Mutational-landscape-SARS-Cov-2

## Results and discussion

We proceed to apply our small-sample support and distribution estimation methods on GISAID SARS-Cov-2 genomic datasets. The underlying assumption is that there exists a “ground truth” distribution of mutations, and that most of the mutations cannot be observed due to limited testing. Our studies of the mutational support and mutation distribution are conducted for different patient subpopulations and all ORFs separately in order to determine potential subpopulation differences. As already pointed out, the estimators to be used are additive implying that estimates for individual genes may be summed to obtain the estimate for the whole genome.

First, we observe that by the last small-sample data collection date reported in the manuscript, 04-14-2020, the average number of mutations with respect to the reference was 7.93 (for male patients) and 7.96 (for female patients). This difference is statistically insignificant. For patients older than 55 years, this number was 7.33 while for those younger than 55 the recorded values were significantly higher, amounting to 8.377. For three different continents, Asia, Europe and NA the average number of mutations recorded equaled 13.51, 6.67, and 6.68, respectively. The average number of mutations per patient in Asia is almost twice as large as the corresponding numbers in Europe and NA, which is indicative of the fact that the outbreak started in Asia and that the virus may have been present in the population significantly longer than in Europe and NA. In all cases, the total number of recorded mutations across all patients is too small to allow for accurate prediction of the actual mutational support using frequency methods.

### Mutational Support

The first set of results pertains to data collected at a very early stage of the pandemic (04-03-2020) that did not include sufficiently many samples to allow for sample set sizes to be evened out through subsampling. Therefore, for this analysis, all available samples are included, which may create biases due to sample set size differences. The results are listed in Tables 4, 5 and 6. They illustrate the difference in the support estimates for two different age groups, genders and three geographic regions. The nonuniform sample size artifacts do not obscure the most important findings regarding mutation rates in different genes across different age groups, gender and geographic region - the same trends persist even when significantly more samples are used in the analysis, as described next.

**Table 4:**
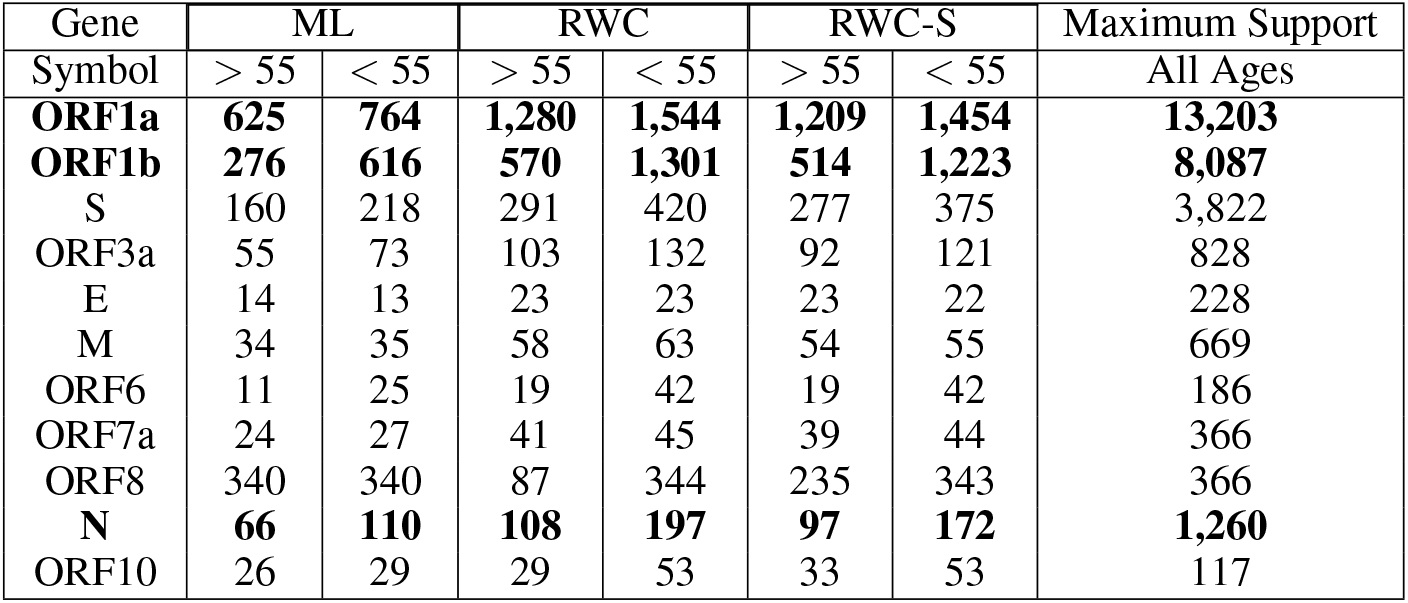
Support sizes of different age groups based on 909 samples for individuals over 55 years of age and 1, 477 samples below 55 years of age. The data was obtained from GISAID by 04-03-2020 and includes all the samples for the two categories available at the given date. ORF1ab and N are shown in **bold** due to their large length and relevance in testing, respectively.

| Gene | ML |  | RWC |  | RWC-S |  | Maximum Support |
| --- | --- | --- | --- | --- | --- | --- | --- |
| Symbol | > 55 | < 55 | > 55 | < 55 | > 55 | < 55 | All Ages |
| **ORF1a** | **625** | **764** | **1,280** | **1,544** | **1,209** | **1,454** | **13,203** |
| **ORF1b** | **276** | **616** | **570** | **1,301** | **514** | **1,223** | **8,087** |
| S | 160 | 218 | 291 | 420 | 277 | 375 | 3,822 |
| ORF3a | 55 | 73 | 103 | 132 | 92 | 121 | 828 |
| E | 14 | 13 | 23 | 23 | 23 | 22 | 228 |
| M | 34 | 35 | 58 | 63 | 54 | 55 | 669 |
| ORF6 | 11 | 25 | 19 | 42 | 19 | 42 | 186 |
| ORF7a | 24 | 27 | 41 | 45 | 39 | 44 | 366 |
| ORF8 | 340 | 340 | 87 | 344 | 235 | 343 | 366 |
| **N** | **66** | **110** | **108** | **197** | **97** | **172** | **1,260** |
| ORF10 | 26 | 29 | 29 | 53 | 33 | 53 | 117 |

**Table 5:** Support sizes based on 1, 349 male and 1, 061 female samples. The data was obtained from GISAID by 04-03-2020 and includes all the samples for the two categories available. ORF1ab and N are shown in **bold** due to their large length and relevance in testing, respectively.

| Gene | ML |  | RWC |  | RWC-S |  | Maximum Support |
| --- | --- | --- | --- | --- | --- | --- | --- |
| Symbol | Male | Female | Male | Female | Male | Female | Both Genders |
| **ORF1a** | **854** | **702** | **1,807** | **1,468** | **1,702** | **1,388** | **13,203** |
| **ORF1b** | **348** | **594** | **690** | **1,307** | **640** | **1,234** | **8,087** |
| S | 225 | 186 | 447 | 359 | 405 | 329 | 3,822 |
| ORF3a | 68 | 61 | 132 | 111 | 115 | 99 | 828 |
| E | 18 | 10 | 30 | 18 | 29 | 18 | 228 |
| M | 37 | 36 | 62 | 68 | 57 | 60 | 669 |
| ORF6 | 13 | 27 | 22 | 49 | 21 | 50 | 186 |
| ORF7a | 32 | 21 | 55 | 38 | 53 | 38 | 366 |
| ORF8 | 340 | 341 | 344 | 592 | 343 | 458 | 366 |
| **N** | **96** | **85** | **165** | **143** | **146** | **129** | **1,260** |
| ORF10 | 26 | 10 | 30 | 17 | 29 | 17 | 117 |

**Table 6:** Support sizes for different geographical regions based on 510 samples from Asia, 1, 695 from Europe and 818 from NA. The data was obtained from GISAID by 04-03-2020 and includes all the samples for the three categories available at the given date. ORF1ab and N are shown in **bold** due to their large length and relevance in testing, respectively.

| Gene | ML |  |  | RWC |  |  | RWC-S |  |  | Maximum Support |
| --- | --- | --- | --- | --- | --- | --- | --- | --- | --- | --- |
| Symbol | Asia | Europe | NA | Asia | Europe | NA | Asia | Europe | NA | All Three Regions |
| **ORF1a** | **770** | **757** | **397** | **1,645** | **1,558** | **776** | **1,603** | **1,455** | **720** | **13,203** |
| **ORF1b** | **279** | **590** | **205** | **566** | **1,251** | **372** | **553** | **1,159** | **345** | **8,087** |
| S | 168 | 181 | 131 | 321 | 345 | 254 | 313 | 309 | 230 | 3,822 |
| ORF3a | 84 | 62 | 38 | 158 | 113 | 71 | 154 | 100 | 63 | 828 |
| E | 37 | 11 | 6 | 66 | 19 | 9 | 65 | 19 | 9 | 228 |
| M | 30 | 29 | 15 | 53 | 49 | 25 | 50 | 44 | 24 | 669 |
| ORF6 | 2 | 28 | 5 | 2 | 46 | 8 | 2 | 45 | 7 | 186 |
| ORF7a | 108 | 38 | 49 | 215 | 66 | 90 | 214 | 65 | 89 | 366 |
| ORF8 | 340 | 27 | 19 | 341 | 46 | 26 | 342 | 43 | 28 | 366 |
| **N** | **53** | **90** | **68** | **93** | **152** | **122** | **85** | **137** | **114** | **1,260** |
| ORF10 | 10 | 25 | 9 | 18 | 28 | 15 | 17 | 27 | 14 | 117 |

Tables 7, 8 and 9 list the results analogue to those reported for 04-03-2020, obtained using datasets retrieved on 04-10-2020. The datasets were sufficiently large to allow for random subsampling to obtain equal sample set sizes for all subpopulations considered (excluding Asia).

**Table 7:** Support sizes of different age groups based on 1, 850 samples from each group. The data was retrieved from GISAID on 04-10-2020. The mutational supports between the two groups differ substantially for the genes shown in *italics*. ORF1ab and N are shown in **bold** due to their large length and relevance in testing, respectively.

| Gene | ML |  | RWC |  | RWC-S |  | Maximum Support |
| --- | --- | --- | --- | --- | --- | --- | --- |
| Name | > 55 | < 55 | > 55 | < 55 | > 55 | < 55 | All Ages |
| **ORF1a** | **996** | **934** | **2,039** | **1,857** | **1,896** | **1,743** | **13,203** |
| **ORF1b** | **499** | **484** | **991** | **965** | **924** | **896** | **8,087** |
| S | 265 | 279 | 490 | 547 | 458 | 501 | 3,822 |
| *ORF3a* | *104* | *79* | *188* | *138* | *171* | *124* | 828 |
| E | 23 | 19 | 36 | 33 | 36 | 32 | 228 |
| M | 55 | 47 | 98 | 86 | 92 | 77 | 669 |
| *ORF6* | *38* | *26* | *65* | *43* | *64* | *41* | *186* |
| *ORF7a* | *60* | *31* | *108* | *50* | *103* | *49* | *366* |
| ORF8 | 340 | 341 | 93 | 342 | 236 | 343 | 366 |
| **N** | **140** | **163** | **248** | **294** | **223** | **265** | **1,260** |
| ORF10 | 31 | 28 | 35 | 49 | 39 | 50 | 117 |

**Table 8:** Support sizes for different genders based on 1, 956 samples for each group. The data was retrieved from GISAID on 04-10-2020. The mutational supports between the two groups differ substantially for the genes shown in *italics*. ORF1ab and N are shown in **bold** due to their large length and relevance in testing, respectively.

| Gene | ML |  | RWC |  | RWC-S |  | Maximum Support |
| --- | --- | --- | --- | --- | --- | --- | --- |
| Name | Male | Female | Male | Female | Male | Female | Both Genders |
| **ORF1a** | **1,071** | **1,115** | **2,176** | **2,313** | **2,055** | **2,175** | **13,203** |
| **ORF1b** | **500** | **804** | **1,013** | **1,721** | **941** | **1,621** | **8,087** |
| S | 283 | 293 | 551 | 562 | 509 | 519 | 3,822 |
| ORF3a | 114 | 99 | 216 | 175 | 190 | 158 | 828 |
| *E* | *24* | *14* | *37* | *23* | *36* | *22* | *228* |
| M | 52 | 56 | 87 | 101 | 82 | 94 | 669 |
| *ORF6* | *42* | *30* | *75* | *51* | *74* | *50* | *186* |
| ORF7a | 42 | 51 | 74 | 87 | 71 | 84 | 366 |
| ORF8 | 341 | 342 | 344 | 345 | 344 | 345 | 366 |
| **N** | **143** | **162** | **251** | **282** | **226** | **259** | **1,260** |
| *ORF10* | *29* | *12* | *33* | *20* | *32* | *19* | *117* |

**Table 9:** Support size for three different geographic regions based on 615 samples from Asia and 1, 000 samples from Europe and NA each. The data was retrieved from GISAID on 04-10-2020. The mutational supports between the three groups differ substantially for the genes shown in *italics*. ORF1ab and N are shown in **bold** due to their large length and relevance in testing, respectively.

| Gene | ML |  |  | RWC |  |  | RWC-S |  |  | Maximum Support |
| --- | --- | --- | --- | --- | --- | --- | --- | --- | --- | --- |
| Name | Asia | Europe | NA | Asia | Europe | NA | Asia | Europe | NA | All Regions |
| **ORF1a** | **827** | **504** | **470** | **1,768** | **975** | **948** | **1,725** | **919** | **874** | **13,203** |
| **ORF1b** | **308** | **271** | **244** | **631** | **531** | **478** | **611** | **491** | **432** | **8,087** |
| S | 182 | 163 | 142 | 352 | 336 | 269 | 340 | 293 | 243 | 3,822 |
| *ORF3a* | *91* | *56* | *39* | *174* | *96* | *74* | *168* | *85* | *63* | *828* |
| *E* | *37* | *12* | *14* | *66* | *21* | *24* | *65* | *21* | *24* | *228* |
| M | 31 | 23 | 17 | 55 | 38 | 28 | 52 | 35 | 27 | 669 |
| *ORF6* | *3* | *48* | *15* | *3* | *87* | *26* | *3* | *86* | *25* | *186* |
| *ORF7a* | *109* | *63* | *51* | *216* | *118* | *94* | *214* | *116* | *93* | *366* |
| ORF8 | 340 | 19 | 21 | 335 | 29 | 31 | 339 | 29 | 31 | 366 |
| **N** | **58** | **72** | **77** | **96** | **121** | **137** | **91** | **108** | **129** | **1,260** |
| *ORF10* | *10* | *26* | *7* | *18* | *48* | *10* | *17* | *48* | *10* | *117* |

Based on the results of Table 7, we see that the mutational supports in populations of different age (cutoff at 55 years) differ substantially for the ORF3a, ORF6 and ORF7a regions (note that ORF1ab and N are shown in **bold** in every table due to their large length and relevance in testing, respectively). For ORF7a, the older population exhibits almost twice as many mutations compared to the younger population, while for ORF6 and ORF3a the corresponding numbers are 1.5 and 1.4, respectively; the estimated mutational supports of the ORF6 and ORF7a regions are close to 1*/*3 of the whole gene length for individuals older than 55 years. The mutational differences in the ORF6 and ORF7a region persist with an increase in the number of samples (see the Supplementary Table S1), with an estimated mutational support for the former region equal almost 1*/*2 of the gene length. Furthermore, additional differences are observed in the M region which were not apparent when using smaller sample set sizes. The protein encoded by ORF6 was studied in depth during the SARS epidemics [41] and it has been established that the ORF6 protein impairs the nuclear import complex formation (controlling the transport of innate immune regulatory cargo to the nucleus of cells capable of increasing antiviral defenses). The protein encoded by ORF7a has been implicated in inhibiting bone marrow stromal antigen 2 virion tethering [42]. Bone marrow stromal antigen 2, also known as tetherin, is an interferon-induced protein which, when expressed, reduces the release of enveloped viral particles. The significant number of predicted mutations in the ORF7a region of older patients suggests a similar observation as that made for the ORF3a region - a possible effort by the virus to disable or strongly impair the function of the tetherin antigen.

The results pertaining to female/male patients differ significantly from those pertaining to different age groups. The results are listed in Table 8, and imply strong differences in the mutation rates of the ORF1b and ORF10 regions. The mutational support of ORF1b in the female population is 1, 621 compared to 941 in the male population, which amounts to a 8.4% difference with respect to the length of the ORF. A similar result is true for the ORF10 region, for which no well-understood functions are known. Some recent results suggest, based on different evidence, that ORF10 encodes a functional protein in SARS-CoV-2 and that positive selection is driving the evolution of this region [43].

The above described differences persist with increased sample set sizes. The estimated mutational support for ORF1b is 24% and 16% of the length of the region, and for ORF10 18% and 32% of the length of the region, for females and males, respectively (see the Supplementary Table S2). Smaller, yet possibly relevant differences are also observed for the ORF3a and M regions, but these do not persist with increased sample set sizes.

For samples obtained from Asia, Europe and NA the results show that despite the number of samples for Asia being significant smaller than that of Europe and NA, the predicted mutational support in all regions is significantly higher except for the N and ORF6 genes (with only 3 mutations observed in the ORF6 gene). This is particularly the case for ORF3a and ORF8, where the mutation rates are more than 2 and 10-fold higher in Asian patients, respectively. It is reasonable to assume that these regions are mutated early on in an epidemic and tend to “accumulate” the number of mutations. Also, the significant differences suggest that the epidemic started *significantly* earlier in Asia than Europe and NA. The ORF3a region is known to encode for a protein that activates the NLRP3 inflammasome [44]. ORF3a proteins are activators of pro-IL-1*β* gene transcription and protein maturation that trigger activation of the NLRP3 inflammasome. The inflammasome has a dual role of boosting the host defense and driving pathologic inflammation. Based on our findings, one possible explanation for the high mutation rate in this region in older populations is that the virus trying to disable the host’s immune system and increase its virulence. Recent results show that the ORF8 protein may be acquired from SARS-related coronaviruses present in bats [45], which could explain the large difference in the mutational support through “adaptation” in a human host (for patients in Asia). The increase in the number of samples available for analysis shows that significant differences in the mutation support of the E, M, ORF6, ORF7a and ORF10 regions exist as well.

Supplementary Tables S1, S2 and S3 show the trends of increase for the mutational support with increased sample sizes. For data collected by 04-14-2020, this includes roughly 9, 000 samples. All sample set sizes used are equal (except for Asia, for which the sample set sizes available are significantly smaller), therefore allowing for fair comparisons. Supplementary Table S1 illustrates that when the sample set sizes are equal, no significant differences are observed in the mutational supports of disparate age groups except in the E, ORF6, ORF7a and ORF8 regions. Given that the difference in the number of mutations in the ORF7a regions persists for several data acquisition dates, the finding appears to be a sample-size independent. On the other hand, the significant differences in the number of mutations in the E region is only evident when sufficiently many samples are available. The E region contains the code for the encapsulation protein of viral RNA, in addition to some spike proteins. In older subjects, this region is subjected to a significantly larger number of mutations than in other groups. This may imply that immunity in elder patient may be dependent on generating antibodies for the encapsulation proteins. Clearly, no conclusive explanation is possible based on limited data sets but the results suggest performing further sampling and analysis for this particular ORF in older patients. Although it has been observed that the immune responses of individuals vary significantly due to the initial viral load, physical health, and the hosts microbiome, no definite link between these features and the mutation rates in the above region can be established due to lack of supporting clinical data at GISAID and other Covid-19 data repositories.

Supplementary Table S2 illustrates surprisingly few differences in the mutational supports of male and female patients once a sufficiently large number of samples is available: Exceptions are the ORF1b and ORF10 regions. For different geographic regions, the most significant difference observed pertains to the ORF8 region, where samples from Asia exhibit a roughly one order of magnitude larger number of mutations compared to those for samples sequenced in Europe and NA. There also exists a marked difference in the mutational support of ORF7a between patients from Europe on one side and patients from Asia and NA on the other (i.e., a roughly two-fold difference for Europe and NA).

Ten additional data collection days (starting on 04-03-2020, ending on 04-14-2020) lead to more than twice the samples, and the results for the latter date are shown in Figure 5 along with the standard deviations of the estimators (note that in order to estimate the variance of an estimator, one needs to subsample the data which requires more samples to start with; hence, the standard deviation is only evaluated for all samples available by 04-14-2020). The additional data samples show that the N region of the SARC-Cov-2 genome exhibits a much more significant increase in mutations than could have be predicted from early small-set sample sizes, amounting to roughly an average of 23% of the genome, across populations. This finding is of significance as it suggests that genomic regions used as identifiers for the virus may mutate much faster than predicted based on small preliminary sample set information. Nevertheless, the N1 and N2 regions used as primer targets for RT-PCR testing (the use of region N3 as a primer has been discontinued) appear to be largely unmutated. This is illustrated in the Supplementary Table S11 which lists a total of only 8 mutations observed in these regions in the SARS-Cov-2 genomes of US patients. Similar results for mutations in viral genomes of patients from China are presented in Supplementary Table S12.

**Figure 5:**
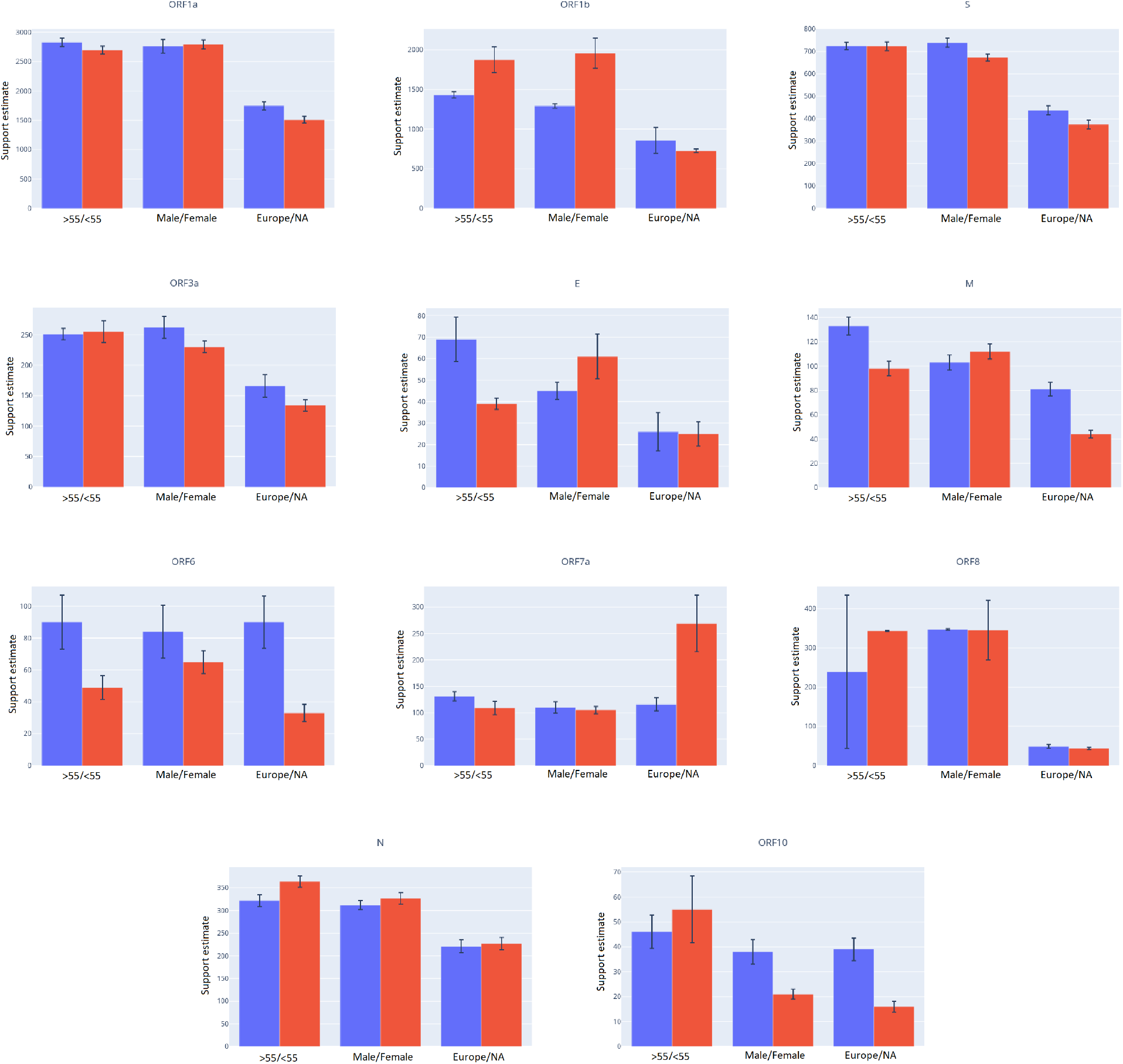
Support sizes for all genes and for different groups along with their standard deviation estimates. The estimates are based on data collected by 04-14-2020 and should provide the most accurate assessment of the mutation rate in the small-sample regimes investigated. Estimates are based on 3, 047 samples each for patients above 55 and below 55 years of age, 2, 817 samples each for male and female patients and 1, 774 each for patients exposed in Europe and North America.

Table 10 provides results for a finer partition of test samples into two categories, one including males over 55 years of age and another females below 55 years of age, with both populations sampled from Europe. The first category has been empirically observed to be at higher risk of infection and for exhibiting more severe symptoms [23]. Substantial mutational differences are observed in the ORF1b, S and ORF10 regions. The differences in the ORF1b and ORF10 genes appear to be mostly gender specific, while the age factor may contribute to the differences in the mutation rates of the S region. Another important finding is that the mutational support of ORF1b is almost twice as large in the low risk population compared to the high risk population. This result may imply that the large mutational support is a result of a highly competitive virus-host interaction which forces the virus to mutate the proteins encoded by ORF1b in order to gain advantage over the host’s immune system.

**Table 10:** Support size differences between males above 55 years of age and females below 55 years of age from Europe based on 1, 078 samples in each group. The data was retrieved from GISAID by 04-14-2020. The mutational supports between the two groups differ substantially for genes with values shown in *italics*. ORF1ab and N are shown in **bold** due to their large length and relevance in testing, respectively.

| Gene | ML |  | RWC |  | RWC-S |  | Maximum Support |
| --- | --- | --- | --- | --- | --- | --- | --- |
| Symbol | M, > 55 | F, < 55 | M, > 55 | F, < 55 | M, > 55 | F, < 55 | Both Categories |
| **ORF1a** | **588** | **670** | **1,159** | **1,374** | **1,078** | **1,294** | **13,203** |
| **ORF1b** | **349** | **553** | **686** | **1,189** | **638** | **1,117** | **8,087** |
| *S* | *209* | *166* | *420* | *329* | *387* | *296* | *3,822* |
| ORF3a | 76 | 61 | 138 | 104 | 124 | 96 | 828 |
| E | 10 | 9 | 17 | 15 | 16 | 14 | 228 |
| M | 27 | 33 | 45 | 58 | 40 | 52 | 669 |
| *ORF6* | *15* | *28* | *25* | *47* | *24* | *48* | *186* |
| *ORF7a* | *31* | *23* | *54* | *36* | *52* | *36* | *366* |
| ORF8 | 27 | 28 | 45 | 48 | 43 | 46 | 366 |
| **N** | **110** | **108** | **197** | **199** | **178** | **183** | **1,260** |
| *ORF10* | *27* | *5* | *28* | *7* | *33* | *7* | *117* |

Figure 5 shows the mutational support sizes, along with the standard deviations of the estimates for six different patient categories. Since the true distribution of mutations of various groups of patients is not known, one cannot directly calculate the standard deviation of the support sizes produced by our estimator. To compute the standard deviation, we therefore subsample 85% of the available samples and compute the support size for the returned aggregate mutation profile. Our samples are chosen randomly and uniformly over the whole period of data collection, and for each month of data collection samples are retrieved separately and in proportion to the total number of samples available for that month. Since the number of samples collected and made available during the months of December and January is small, we group these two months together in the subsampling process. Subsampling is repeated 100 times resulting in 100 aggregate mutation profiles and corresponding support size estimates.

The mutational supports generated by our procedure have variances that demonstrate good concentration of the estimates; some exceptions exist, though, and are most likely not a consequence of the estimation procedure itself but rather an indicator of disparately collected datasets or some unknown governing biological process. The latter is supported in part by previously observed high rates of mutations in certain SARS-Cov-2 genes [46, 47]. The results for ORF8 are particularly interesting because the corresponding standard deviations of the mutational support vary significantly across different categories of patients: The standard deviation of the support size is close to 200 times higher for patients above 55 years of age then patients below 55 years of age.

We also performed a collection of tests in which alignments and mutational counts were performed with respect to the first sample from the same geographical region. Hence, for patients from Asia, the alignments and mutation counts are still performed with respect to the genome of the Wuhan-Hu-1 patient. For NA, we used the sample USA/WA1/2020 with ID EPI_ISL_404895, while for Europe we used the sample France/IDF0372/2020 with ID EPI_ISL_406596, both being the chronologically first samples from NA and Europe available at GISAID. For this study, we only used samples retrieved by 04-14-2020. The results are available in Supplementary Tables S4-S7. As expected, the mutational support estimates are lower for both the NA and European sample sets. However, one important and interesting exception pertains to the estimates for the gene N regions and samples from Europe, as well as samples for males above 55 from Europe, which are higher for the alignment and mutation counts performed with respect to Patient 1 in Europe. The same is true for mutational support estimates for gene N and under gender stratification. Additional differences were observed in the mutational support of the ORF6a and ORF7 regions in younger females versus older males when focusing on patients from Europe only and when using Patient 1 from Europe as the alignment reference. These results suggest different mutational patterns for viruses hosted by high-risk populations in Europe versus those in NA and Asia.

It is important to note that for some genes and patient categories it appears the RWC estimates roughly double those ML estimator but this is **not a general trend** of the analysis. For example, the mutational support estimates for ORF8 for male and female are approximately equal to ML estimates (Table 8) and more pronounced differences exist across the whole subpopulation spectrum. Similar trends are observed for ORF6 in Asian subjects, and ORF10 across different subpopulations. Furthermore, although **the naive ML estimates may lead to similar conclusions regarding the trends of mutations in some ORFs, the degree of the trend and the scale of the mutation rates within different regions cannot be fully understood through the use of ML estimates only**. As an illustrative example, the ML estimator implies that there is no difference in the mutational supports of the ORF8 region in young versus old patients (Table 7), as the values equal 340 and 341, respectively. On the other hand, the RWC-S estimator predicts mutational supports of 236 and 343, respectively, which show a very different stratification.

We conclude by pointing out that one way to validate the results for our support estimation methods is to compare the results of the ML mutation counts at a later date with the computed estimates. We compare the mutational supports using the small-sample techniques and the data collected by 04-10-2020 with the actual count (ML estimates) generated from data retrieved by 04-14-2020. In this time period, the number of samples increased by roughly 3, 000, as may be seen from Table 1. The results are listed in Table 11. As may be seen, the estimates obtained based on data acquired by 04-10-2020 for Europe and NA and all open reading frames are excellent matches for the actual counts obtained by 04-14-2020, indicating that the number of samples was sufficient to predict the growth trend. Much more significant differences are observed for Asia, which can clearly be attributed to the very small-sample sizes available from that continent on both 04-10-2020 and 04-14-2020 or potential strong correlations between the mutations in the three aforementioned regions. Other categories that are of interest involve male/female patients for which the actual counts from 04-14-2020 are significantly smaller than the estimates. This is indicative of a large number of potentially unseen mutations harbored by these populations.

**Table 11:** Comparison of small-sample estimates of RWC-S based on data retrieved by 04-10-2020 and the ML estimates based on data retrieved by 04-14-2020.

| Gene | Method and Date | Asia | Europe | NA | Above 55 | Below 55 | Male | Female |
| --- | --- | --- | --- | --- | --- | --- | --- | --- |
| ORF1a | RWC-S (04-10-2020) | 1,725 | 919 | 874 | 1,896 | 1,743 | 2,055 | 2,175 |
|  | ML (04-14-2020) | 835 | 911 | 804 | 1,488 | 1,439 | 1,478 | 1,456 |
| ORF1b | RWC-S (04-10-2020) | 611 | 491 | 432 | 924 | 896 | 941 | 1,621 |
|  | ML (04-14-2020) | 316 | 477 | 403 | 787 | 953 | 705 | 991 |
| S | RWC-S (04-10-2020) | 340 | 293 | 243 | 458 | 501 | 509 | 519 |
|  | ML (04-14-2020) | 188 | 246 | 209 | 431 | 400 | 405 | 389 |
| ORF3a | RWC-S (04-10-2020) | 168 | 85 | 63 | 171 | 124 | 190 | 158 |
|  | ML (04-14-2020) | 93 | 99 | 81 | 156 | 165 | 169 | 140 |
| E | RWC-S (04-10-2020) | 65 | 21 | 24 | 36 | 32 | 36 | 22 |
|  | ML (04-14-2020) | 36 | 15 | 15 | 43 | 26 | 30 | 36 |
| M | RWC-S (04-10-2020) | 52 | 35 | 27 | 92 | 77 | 82 | 94 |
|  | ML (04-14-2020) | 31 | 51 | 28 | 79 | 62 | 67 | 69 |
| ORF6 | RWC-S (04-10-2020) | 3 | 86 | 25 | 64 | 41 | 74 | 50 |
|  | ML (04-14-2020) | 3 | 52 | 21 | 53 | 32 | 50 | 40 |
| ORF7a | RWC-S (04-10-2020) | 214 | 116 | 93 | 103 | 49 | 71 | 84 |
|  | ML (04-14-2020) | 109 | 66 | 135 | 86 | 66 | 68 | 72 |
| ORF8 | RWC-S (04-10-2020) | 339 | 29 | 31 | 236 | 343 | 344 | 345 |
|  | ML (04-14-2020) | 340 | 32 | 29 | 341 | 343 | 343 | 342 |
| N | RWC-S (04-10-2020) | 91 | 108 | 129 | 223 | 265 | 226 | 259 |
|  | ML (04-14-2020) | 60 | 139 | 138 | 201 | 219 | 195 | 204 |
| ORF10 | RWC-S (04-10-2020) | 17 | 48 | 10 | 39 | 50 | 32 | 19 |
|  | ML (04-14-2020) | 11 | 30 | 10 | 35 | 33 | 31 | 13 |

Finally, Table 12 shows the support estimates for samples from patients from Asia for a more recent date of data collection, 10-20-2020. In this case, almost 10, 000 samples from Asia were readily available which allows one to get significantly improved results for ML estimators. As may be seen, the differences between ML and RWC-S values are significantly smaller, and for some reasons even close to equal when a very different trend was true for data collected in April. In particular, the ratio of the number of estimated mutations in the ORF E region for the RWC-S and ML method was close to 1.76 in April, and only 1.24 in October. Similar findings are apparent for other ORFs.

**Table 12:** ML and RWC-S estimates for mutational support in ORFs of patients from Asia based on data collected by 10-20-2020. At that date, substantially more samples (9, 271) were available for analysis. The standard deviation values are given in parentheses.

|  | ORF1a | ORF1b | S | ORF3a | E | M | ORF6 | ORF7a | ORF8 | N | ORF10 |
| --- | --- | --- | --- | --- | --- | --- | --- | --- | --- | --- | --- |
| ML | 5,020<br>(77) | 2,691<br>(42) | 1,418<br>(27) | 464<br>(19) | 115<br>(3) | 188<br>(4) | 90<br>(3) | 304<br>(9) | 363<br>(1) | 510<br>(6) | 45<br>(2) |
| RWC-S | 8,481<br>(152) | 4,435<br>(80) | 2,227<br>(61) | 674<br>(34) | 143<br>(5) | 262<br>(8) | 112<br>(7) | 333<br>(15) | 361<br>(7) | 711<br>(14) | 61<br>(3) |

**Table 13:** The Jaccard distance between sets of mutations from different pairs of geographic regions, based on alignments with respect to Patient 1 from Wuhan. Values in *italics* are the smallest in the category, while values in **bold** are the largest.

|  | ORF1a | ORF1b | S | ORF3a | E | M | ORF6 | ORF7a | ORF8 | N | ORF10 |
| --- | --- | --- | --- | --- | --- | --- | --- | --- | --- | --- | --- |
| Asia - Europe | **0.91** | **0.95** | 0.91 | 0.92 | **0.98** | **0.92** | **0.96** | *0.74* | 0.91 | 0.86 | *0.89* |
| Europe-NA | 0.88 | 0.89 | 0.88 | 0.84 | 0.89 | 0.89 | *0.86* | **0.91** | 0.89 | 0.82 | 0.95 |
| Asia - NA | **0.91** | **0.95** | **0.92** | **0.95** | 0.91 | 0.89 | **0.96** | 0.89 | **0.92** | **0.87** | *0.89* |
| Male >55 - Female <55 (Europe) | *0.85* | *0.87* | *0.85* | *0.73* | *0.88* | *0.75* | 0.87 | *0.83* | *0.83* | *0.77* | **0.97** |

### Distribution Estimation

Next, we report on the distribution of mutations in the ORF1a,b and N regions of the SARS-Cov-2 virus obtained using the Good-Turing estimator and once again focus on the traits of different subpopulations. We focus on these regions as the first two regions are the longest genes while the N region is of importance for Covid-19 testing in NA. As may be seen from Figures 6 and 7 there is a surprisingly small difference in the distribution of the top-20 mutated sites across different age and gender groups, except for a marked difference in the largest probability (in particular, in the N region for populations partitioned according to age and populations partitioned according to gender when including larger sample sets from 04-14-2020 as seen in Supplementary Figure S1). This is especially the case for samples partitioned according to gender, despite the fact that the number of mutations in female subjects in the ORF1b region was close to twice as large as that in male subjects. In addition, the probability of having a mutation at the highest probability sites is significantly larger in “younger” than “older” populations. The trend remains the same for data collected by 04-14-2020 as supported by the results in Supplementary Figure S1. Supplementary Figure S2 gives similar results for alignment performed against first sequenced patient from each region. The situation is completely different when comparing the distributions of mutations across different geographic regions (Figure 8), where there are significant differences in the distributions as one would expect. To compactly summarize the differences in the distributions, we also computed all three pairwise symmetric Kulback-Leibler (KL) divergences for the normalized top-20 mutation probabilities as described below. We also list the Jaccard distances between the sets of 20 most frequently mutated sites.

**Figure 6:**
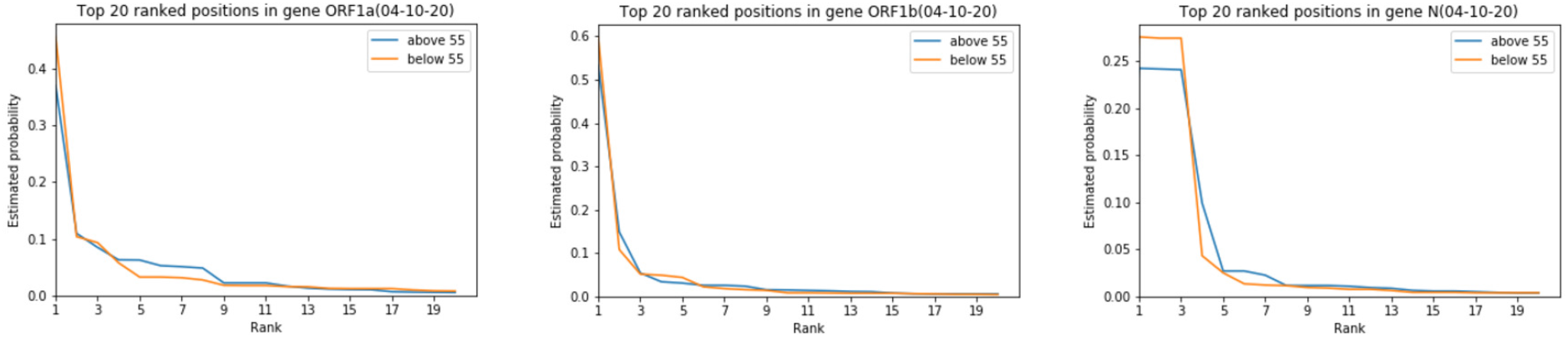
Comparison of the estimated distributions of mutations for genes ORF1a, ORF1b and N in adults *<*55 of age and adults *>*55 of age tested by 04-10-2020. Almost all the probability mass is concentrated on five sites. The biggest observed difference occurs in the N region.

**Figure 7:**
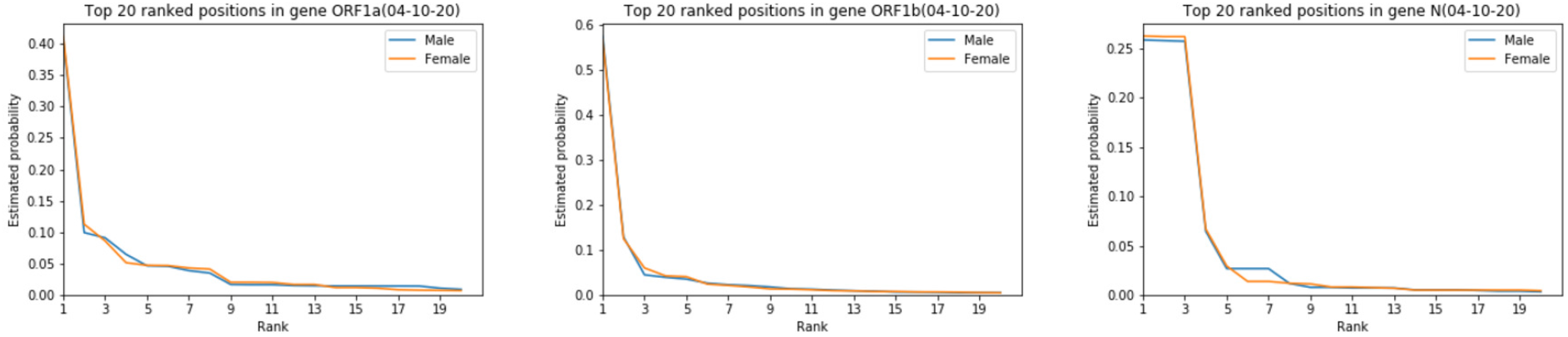
Comparison of the estimated distributions of mutations for genes ORF1a, ORF1b and N in male and female test subjects tested by 04-10-2020. The distributions exhibit no difference except on two sites in the N region.

**Figure 8:**
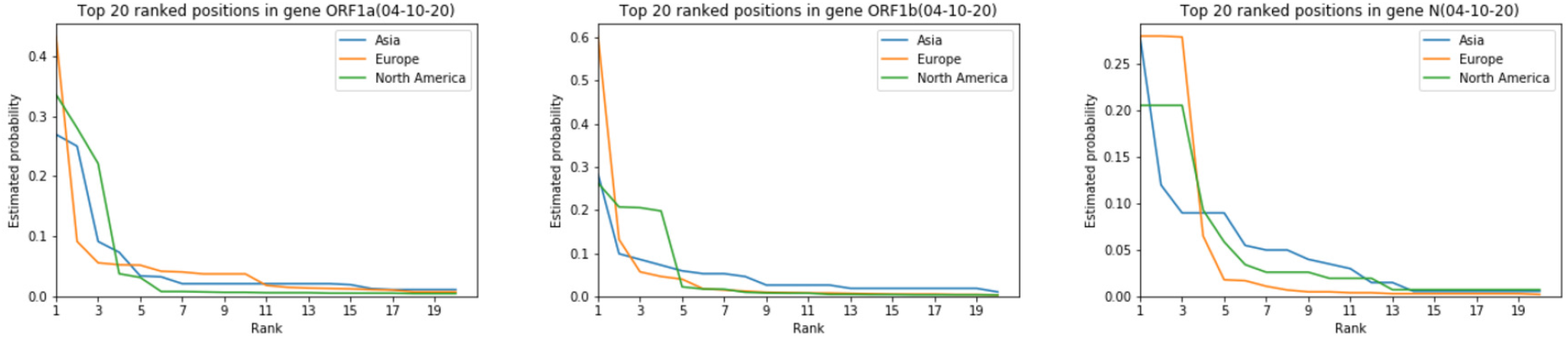
Differences in the estimated distributions of mutations for genes ORF1a, ORF1b and N for different geographic regions based on subjects tested by 04-10-2020. The distributions differ significantly.

The distributions of mutations only reveal the statistical landscape of the mutation sites but not their exact locations in the genome. The actual mutated sites in the SARS-Cov-2 genomes are depicted in Figures 9, 10, in addition to a more detailed set of results presented in the Supplementary Figures S3 and S4. We selected the latest retrieval data for this analysis as it most accurately reflects the positions undergoing most frequent mutations; we also focused on two cohorts of patients for which the mutational landscapes differ the most. The positional stratification of mutations is significant for patients from different continents, especially in the N region of the SARS-Cov-2 genome. The largest spread of probability mass is (as expected) observed for patients from Asia which is indicative of the larger exploration rate for mutations in the region where the outbreak originated.

**Figure 9:**
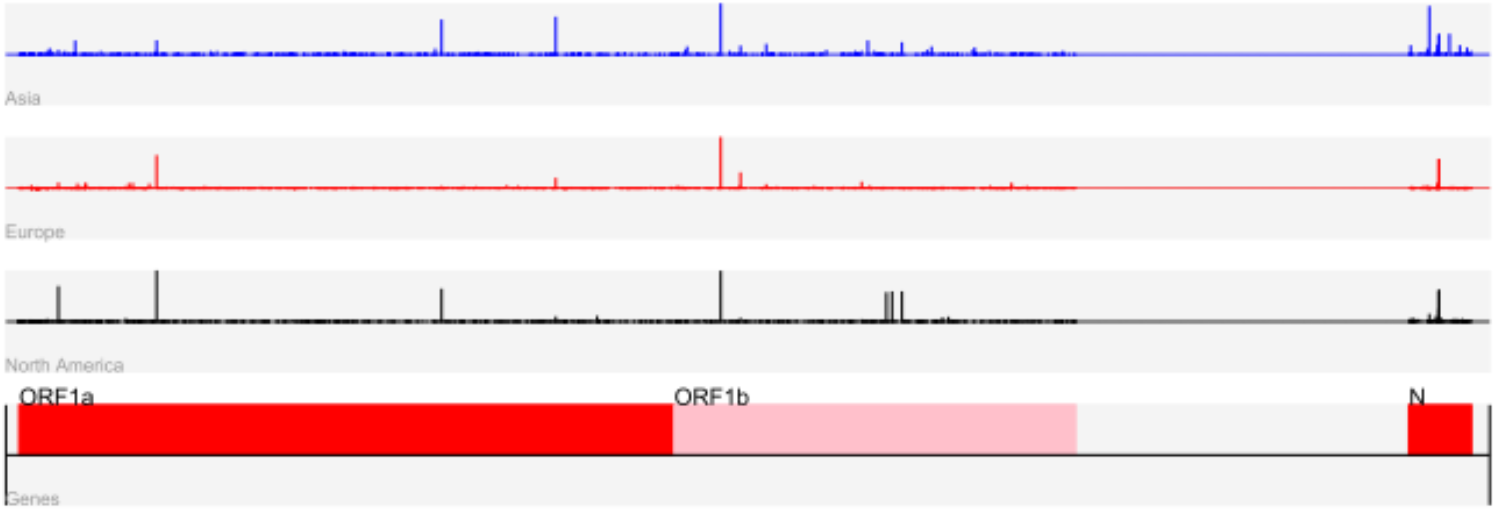
Positions of mutations in the SARS-Cov-2 genome for patients across three different continents, for data collected by 04-14-2020. The height of the bar is proportional to the estimated probability of mutation.

**Figure 10:**
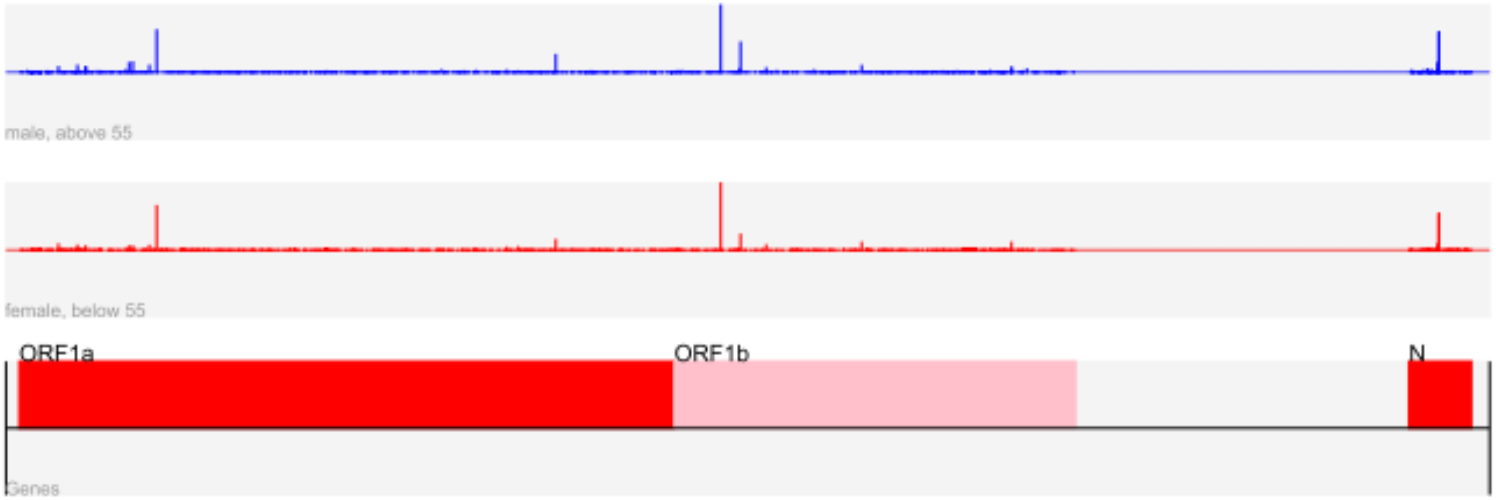
Positions of mutations in the SARS-Cov-2 genome for European females of age *<* 55 and males of age *>* 55 collected by 04-14-2020. The height of the bar is proportional to the estimated probability.

Supplementary Table S8 lists the 10 most frequently mutated sites in the ORF1a region of all previously analyzed patients categories when alignment is performed with respect to the first patient sequenced in the geographic region. For the age and gender groupings, as expected, the top-ten sites are the same except for one difference encountered in both cases (shown in **bold**). A mutation in position 8, 781 of Asian and NA viral samples appears with high frequency but is surprisingly not present in the list of top mutated sites in the European population. Similarly, Supplementary Table S9 lists the 10 most frequently mutated sites in the ORF1b region of all previously analyzed patients categories. As one may expect from the differences in the mutational support, the frequent sites of mutations differ significantly more in this region for different age groups, gender and continents when compared to the ORF1a region. This is especially the case when viewing the results for different geographic regions as except for the top-ranked site and one more site (i.e., sites 14, 407 and 14, 804), all other locations are different. This suggests very different evolution patterns of the virus in the ORF1b genomic region at different regional sites, and more similar mutational patterns for different gender and age categories. Supplementary Table S10 suggests significantly fewer stratifications in the mutations of different patient groups in the N region. Gender and age does not appear to play a major role, but marked differences are observed in patients from Asia, Europe and NA (the sites mutated in two regions but not in the third are shown in *italics*). Given the large differences in the mutational sites of patients across different continents in the N region it does not come as a surprise that different primers for RT-PCR testing were selected for Asia, Europe and NA. The sites selected for forward and reverse primers by the CDC, i.e., the N1 and N2 region, do not contain a significant number of mutations, as may be easily seen from Supplementary Table S11. Similar observations are true for the primers selected in China (Supplementary Table S12).

The outlined distribution estimation procedure may also be important in terms of predicting growth trends for certain mutations that may not be easily observed based on frequency counts alone. Figure 11 demonstrates that the Good-Turing estimator predicted higher probabilities for 4 out of 6 mutation sites in the S region of the UK variant compared to the ML estimator for data collected in September 2020, giving a potentially early indication of the spread of the UK variant (when very few samples with the underlying mutations are seen). Using data from November 2020 in conjunction with the GT and ML estimators produced identical results.

**Figure 11:**
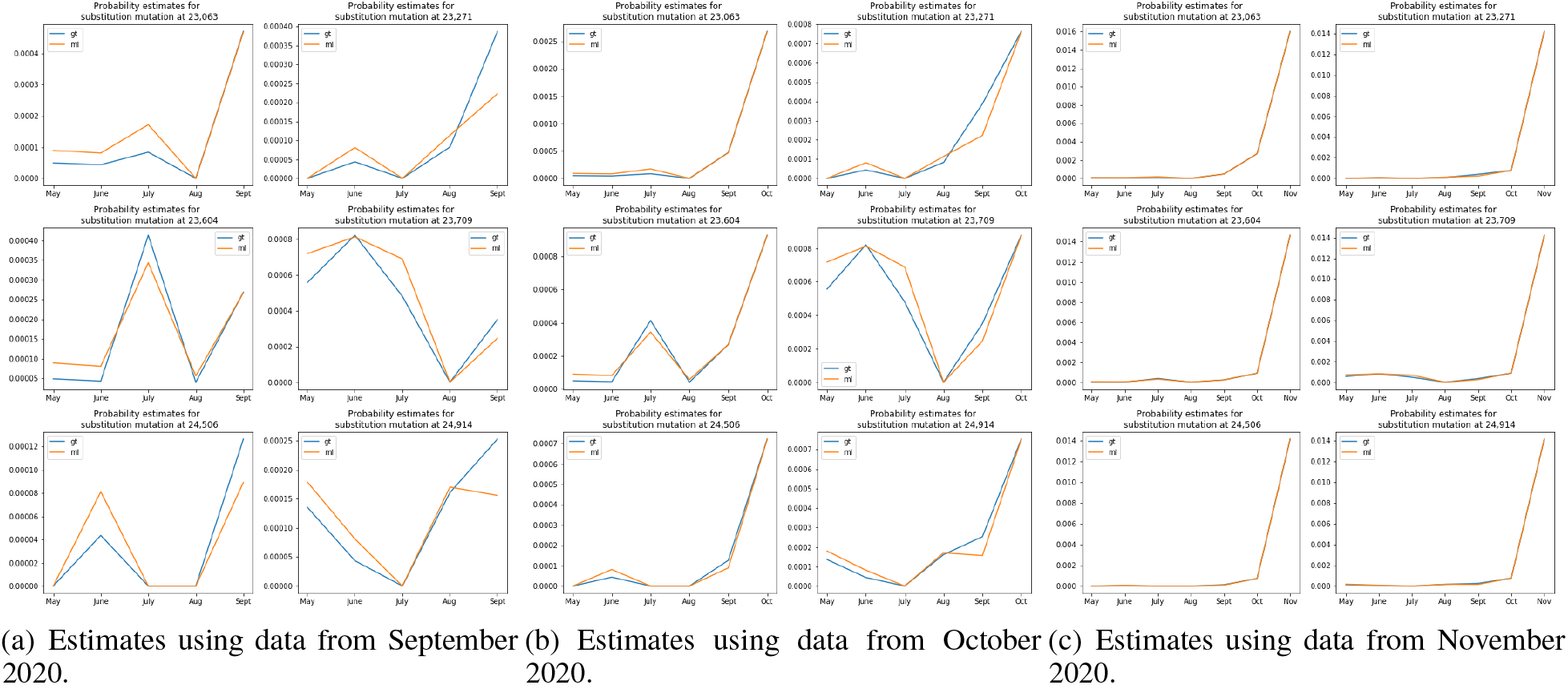
The probability of mutations at six substitution sites in the S region of the UK variant obtained using the GT and ML estimators.

### Summarizing the Differences in the Distributions Using the Symmetric KL Divergence and the Jaccard Distance

The symmetric KL divergence between two discrete probability distributions *p* and *q* is defined as

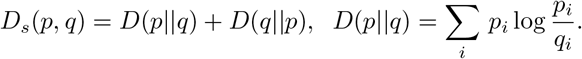

For the mutation distributions pertaining to Europe-NA, Europe-Asia and Asia-NA, the KL divergences equal 0.672, 0.316 and 0.376 (ORF1a), 0.491, 0.435 and 0.646 (ORF1b), 0.293, 1.021 and 0.303 (N), respectively, for data collected by 04-14-2020. These results indicate that the largest differences in the distributions in the ORF1a region exist between Europe and NA, while the largest differences in the ORF1b region exist between Asia and NA. For the N region, a significant difference between the distributions of mutations is observed between Europe and Asia, and at this point, no simple explanation for this finding is possible. Similarly, the corresponding KL divergences based on the samples collected by 04-10-2020 equal 0.788 (which is significantly larger than the one predicted based on data collected on 04-14-2020), 0.328 and 0.371 (ORF1a), 0.743 (which is significantly larger than the one predicted based on data collected on 04-14-2020), 0.615 and 0.0.755 (ORF1b), 0.315, 0.893 and 0.248 (N), respectively. The results for the KL divergences for the N regions suggest relatively small changes in the distribution of mutations in the N region, and larger changes in the ORF1a and ORF1b regions, which is expected.

Since the previously described distribution estimates do not convey the information about the locations of the highest mutated sites but only their frequency of mutations, we also list the Jaccard distances of the sets of mutations specific to each tested subpopulation. For two sets Σ_1_ and Σ_2_ over the same ground set Σ, the Jaccard distance *J*(Σ_1_, Σ_2_) is defined as:

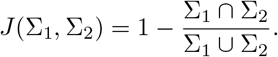

As may be seen from Table 13, the largest distances are observed in the E and ORF10 regions, in the first case when comparing patients from Asia and Europe and in the second case when comparing younger female and older males in Europe. The distances in the N region seem to be significantly smaller, especially between the two categories of patients from Europe. The results for the ORF10 region are rather surprising as they indicate the highest possible difference is observed between males and females on the same continent despite these differences being uniformly small for all other open reading frames. As already pointed out, the function of the ORF10 reading frame is currently unknown but given the marked mutational profiles in high-risk and low-risk profiles it is highly likely that this gene plays an important role in guiding disease symptoms. The exact same trends are observed when using alignments with respect to Patient 1 from the underlying geographic region, as listed in Supplementary Table 13.

## Conclusion

The problem of determining mutational support and distribution of a virus is crucial for accessing its virulence and for primer selection for real time RT-PCR kits, especially during early stages of a pandemic when insufficient amount of information is available about the virus. An accurate estimate of possible mutations in the viral genome in the absence of a sufficiently large database is important for an early understanding of the adaptation mechanisms employed by the virus as well as the potential differences in its impact on diverse subpopulations. In this paper, we presented a novel, state-of-the-art estimator for support estimation for the small-sample regime and benchmarked it against existing estimators. We also adapted the Good-Turing estimator for distribution estimation.

We used our estimators for a differential analysis on mutations in the SARS-Cov-2 genome among various population groups including male/female, older/younger and different geographic locations. We observed significant differences in the mutational support of ORF6 and ORF7a between older and younger patients as well as differences in ORF1b and ORF10 between males and females. We also noted that these differences persist with increase in number of samples available. Given that these ORFs play important biological roles in the spread and evolution of the virus, these differences can provide significant insights into why different population groups are impacted differently by the virus. Furthermore, we discovered differences in mutational support among all ORFs while comparing between different geographical locations. Our analysis showed that patients from Asia had comparatively higher mutational support than those from Europe and North America, which can potentially indicate that the virus was in circulation much earlier than expected. We validated our results by comparing the support estimate returned by our estimators on 04-10-2020 with ML estimates from 04-14-2020 as well as comparing the two estimators on a much larger sample set obtained on 10-20-2020.

We observed that even though the N region of SARS-CoV-2 genome has a high number of mutations, only a few mutations lay in the primer regions for real time RT-PCR kits recommended by CDC for testing in the USA. This is important because frequent mutations in the primer regions can potentially lead to high rates false negative results. Finally, we compared the distributions of mutations among various population groups and compute pairwise symmetric Kulback-Leibler divergences for normalized top-20 mutated positions as well as Jaccard distance for the sets of all mutations for each population. We would emphasize that our estimators are general enough to be adapted for the genomes of any microorganism making it extremely useful in the early stages for any future outbreak as well.

## Data Availability

All sequencing data we used is available from GISAID under hCoV-19

https://www.gisaid.org/

## Supporting information

**S1 Text. Proofs and additional theoretical results**. Proofs of various theorems presented in the paper and additional results corresponding to the algorithms developed.

**S1 Fig. Distribution of mutations**. Comparison of mutations in various groups of patients based on the data was collected by 04-14-2020. All the alignments were performed with respect to Patient 1 Wuhan-Hu-1.

**S2 Fig. Distribution of mutations**. Comparison of mutations in various groups of patients based on the data was collected by 04-14-2020. All the alignments were performed with respect to the first sequenced patient 1 in the corresponding region.

**S3 Fig. Position of mutations**. Positions of mutations in the SARS-Cov-2 genome with high probability of mutations in patients from different categories based on data collected by 04-14-2020. The height of the bar is proportional to the probability of the mutation. All the alignments were performed with respect to Patient 1 Wuhan-Hu-1.

**S4 Fig. Position of mutations**. Positions of mutations in the SARS-Cov-2 genome with high probability of mutations in patients from different categories based on data collected by 04-14-2020. The height of the bar is proportional to the probability of the mutation. All the alignments were performed with respect to the first sequenced patient 1 in the corresponding region.

**S1-S3 Table. Support estimate**. Support sizes for all the genes for different groups of patients based on data collected by 04-14-2020. All the alignments were performed with respect to Patient 1 Wuhan-Hu-1.

**S4-S7 Table. Support estimate**. Support sizes for all the genes for different groups of patients based on data collected by 04-14-2020. All the alignments were performed with respect to the first sequenced patient 1 in the corresponding region.

**S8-S10 Table. Frequently mutated sites**. Locations of 10 most frequently mutated positions for different groups of patients for genes ORF1ab and N.

**S11 Table. Mutations in primer region (USA)**. The total number of mutations across all samples of patients from the USA in positions corresponding to primers used in the corresponding RT-PCR kits. The positions are along the length of the reference genome published in (JHU). The number of samples is 1, 764 collected by 04-14-2020.

**S12 Table. Mutations in primer region (China)**.The total number of mutations across all samples from patients in China, at positions corresponding to the primers used in their corresponding RT-PCR kits. The positions are along the length of the reference genome published in (JHU). The number of samples is 395, with collection date 04-14-2020.

**S13 Table. Jaccard distance**.The Jaccard distance between sets of mutations from different pairs of geographic regions. The results are based on alignments with respect to Patient 1 from the corresponding geographic region.

