## Supplementary material for "Small-sample estimation of the mutational support and the distribution of mutations in the SARS-Cov-2 genome": Supp Text

#### 1 Proofs

##### 1.1 Proof of the Theorem Establishing the Length of the Optimization Interval

To prove the result, we need to show that $\forall \lambda \geq 6.5L$, $\frac{\partial}{\partial \lambda} g(\mathbf{a}, \lambda) < 0$. The derivative of the first term in $g$ equals

$$\frac{\partial}{\partial \lambda} \frac{1}{k} \Big( \sum_{l=0}^{L} e^{-\lambda} a_l^2 \lambda^l l! \Big) = \frac{1}{k} \Big( \sum_{l=0}^{L} (\frac{l}{\lambda} - 1) e^{-\lambda} a_l^2 \lambda^l l! \Big).$$

Clearly, the right hand side in the above expression is negative for all $\lambda > L$. The second term of the derivative equals

$$\begin{aligned}
&\frac{\partial}{\partial \lambda} \Big( e^{-\lambda} \sum_{l=0}^{L} a_l \lambda^l \Big)^2 \\
&= 2 \Big( e^{-\lambda} \sum_{l=0}^{L} a_l \lambda^l \Big) \Big( - e^{-\lambda} \sum_{l=0}^{L} a_l \lambda^l + e^{-\lambda} \sum_{l=0}^{L} \frac{l}{\lambda} a_l \lambda^l \Big) \\
&= 2 e^{-2\lambda} \Big( \sum_{l=0}^{L} a_l \lambda^l \Big) \Big( \sum_{l=0}^{L} (\frac{l}{\lambda} - 1) a_l \lambda^l \Big).
\end{aligned}$$

To analyze the two terms of the derivative, we introduce the vectors $\mathbf{y}, \mathbf{z}, \mathbf{1}$ and the diagonal matrix $\mathbf{D}$ according to

$$\begin{aligned}
&\mathbf{y} = (a_0 \lambda^0, a_1 \lambda^1, ..., a_L \lambda^L)^T, \\
&\mathbf{z} = ((\frac{0}{\lambda} - 1), (\frac{1}{\lambda} - 1), ..., (\frac{L}{\lambda} - 1))^T, \\
&\mathbf{1} = (1, 1, ..., 1)^T, \\
&D_{ii} = (-1 + \frac{i-1}{\lambda}) \frac{(i-1)!}{\lambda^{(i-1)}}.
\end{aligned}$$

Consequently, we have

$$\begin{aligned}
&\frac{\partial}{\partial \lambda} \frac{1}{k} \Big( \sum_{l=0}^{L} e^{-\lambda} a_l^2 \lambda^l l! \Big) = \frac{e^{-\lambda}}{k} \mathbf{y}^T \mathbf{D} \mathbf{y}, \\
&\frac{\partial}{\partial \lambda} \Big( e^{-\lambda} \sum_{l=0}^{L} a_l \lambda^l \Big)^2 = 2 e^{-2\lambda} \mathbf{y}^T \mathbf{1} \mathbf{z}^T \mathbf{y} = e^{-2\lambda} \mathbf{y}^T (\mathbf{1} \mathbf{z}^T + \mathbf{z} \mathbf{1}^T) \mathbf{y}.
\end{aligned}$$

Therefore,

$$\frac{\partial}{\partial \lambda} g(\mathbf{a}, \lambda) = e^{-2\lambda} \mathbf{y}^T \left( \frac{e^{\lambda}}{k} \mathbf{D} + (\mathbf{1} \mathbf{z}^T + \mathbf{z} \mathbf{1}^T) \right) .\mathbf{y}$$

To show that $\frac{\partial}{\partial \lambda} g(\mathbf{a}, \lambda) < 0$ for all polynomials of degree $L$ whenever $\lambda > CL$, we show that the matrix $\left( \frac{e^{\lambda}}{k} \mathbf{D} + (\mathbf{1} \mathbf{z}^T + \mathbf{z} \mathbf{1}^T) \right)$ is negative-definite whenever $\lambda > CL$, for some constant $C > 0$. It suffices to show that the sum of the maximum eigenvalues of $\frac{e^{\lambda}}{k} \mathbf{D}$ and $(\mathbf{1} \mathbf{z}^T + \mathbf{z} \mathbf{1}^T)$ is negative, since $\frac{e^{\lambda}}{k} \mathbf{D}$ is a diagonal matrix. Thus, we turn our attention to determining the maximum eigenvalues of these two matrices. For $\frac{e^{\lambda}}{k} \mathbf{D}$, the maximum eigenvalue satisfies

$$\frac{e^{\lambda}}{k} \max_{i \in \{0,1,...,L\}} \left( -1 + \frac{i}{\lambda} \right) \frac{i!}{\lambda^i} \leq -\frac{e^{\lambda}}{2k} \min_{i \in \{0,1,...,L\}} \frac{i!}{\lambda^i},$$

since for $\lambda > 2L$, one has $(-1 + \frac{i}{\lambda}) \leq -\frac{1}{2}$. When $\lambda > L$, it is clear that $\frac{i!}{\lambda^i}$ is decreasing in $i$, for $i \in \{0, 1, ..., L\}$, so that

$$\min_{i \in \{0,1,\ldots,L\}} \frac{i!}{\lambda^i} = \frac{L!}{\lambda^L} \geq \left(\frac{L}{e\lambda}\right)^L.$$

The last inequality is a consequence of Stirling's formula, which asserts that $n! \geq (\frac{n}{e})^n$. Combining the above expressions, we obtain

$$\frac{e^\lambda}{k} \max_{i \in \{0,1,\ldots,L\}} \left(-1 + \frac{i}{\lambda}\right) \frac{i!}{\lambda^i} \leq -\frac{e^\lambda}{2k} \left(\frac{L}{e\lambda}\right)^L.$$

Next, we derive an upper bound on maximum eigenvalue of the second matrix. The $i, j$ entry of the matrix $(\mathbf{1}\mathbf{z}^T + \mathbf{z}\mathbf{1}^T)$ equals $\frac{i+j-2}{\lambda} - 2$, and all these values are negative when $\lambda > L$. Moreover, it is clear that the matrix of interest has rank equal to 2. Therefore, the matrix has exactly two nonzero eigenvalues.

Let $\mathbf{A} = -(\mathbf{1}\mathbf{z}^T + \mathbf{z}\mathbf{1}^T)$. All entries of $\mathbf{A}$ are positive whenever $\lambda > L$. By Gershgorin's theorem, we can upper bound the maximum eigenvalues of the matrix $\mathbf{A}$ by its maximum row sum. It is obvious that the maximum row sum equals

$$2(L+1) - \frac{L(L+1)}{2\lambda}.$$

Moreover, the trace of $\mathbf{A}$ equals

$$2(L+1) - \frac{L(L+1)}{\lambda}.$$

This implies that the minimum eigenvalue of $\mathbf{A}$ is lower bounded by $-\frac{L(L+1)}{2\lambda}$, which directly implies that the maximum eigenvalue of $(\mathbf{1}\mathbf{z}^T + \mathbf{z}\mathbf{1}^T)$ is upper bounded by $\frac{L(L+1)}{2\lambda}$.

Summing up the two previously derived upper bounds gives

$$h(\lambda) \triangleq -\frac{e^\lambda}{2k} \left(\frac{L}{e\lambda}\right)^L + \frac{L(L+1)}{2\lambda},$$

whenever $\lambda > 2L$. Note that $h(\lambda) < 0$ is equivalent to

$$\frac{L(L+1)}{2\lambda} < \frac{e^\lambda}{2k} \left(\frac{L}{e\lambda}\right)^L$$
$$\Leftrightarrow \log(L) + \log(L+1) + \log(k) - L\log(L) + L < \lambda + \log(\lambda) - L\log(\lambda). \quad (1)$$

The function $\lambda + \log(\lambda) - L\log(\lambda)$ is nondecreasing in $\lambda$ whenever $\lambda > L$ since

$$\frac{d}{d\lambda}(\lambda + \log(\lambda) - L\log(\lambda)) = 1 - \frac{L-1}{\lambda}.$$

By the definition of $L = \lfloor c_0 \log(k) \rfloor$, we also have $\log(k) \leq \frac{L+1}{c_0}$. Using $\log(x+1) \leq x$, which holds $\forall x \geq 1$. Hence $\forall \lambda > CL$ where $C > 2$, the sufficient condition for (1) to hold is

$$\log(L) + L + \frac{L+1}{c_0} - L\log(L) + L < CL + \log(CL) - L\log(CL).$$

Rearranging terms leads to

$$\left(C - \log(C) - 2 - \frac{1}{c_0}\right) L + \log(C) > \frac{1}{c_0}.$$

Sufficient conditions that ensure that the above inequality holds are $\log(C) \geq \frac{1}{c_0}$ and $(C - \log(C) - 2 - \frac{1}{c_0}) > 0$. The first condition implies $C \geq e^{\frac{1}{c_0}} = 6.0021$, while the second condition holds with $C = 6.5$, for which the first condition is also satisfied. This completes the proof.

#### 1.2 Proof of the convergence rate of the discretized SIP

The proof consists of two parts. In the first part, we establish the conditions for convergence, while in the second part, we determine the convergence rate. For simplicity, we present the proofs for the case without Poisson repeats. We then outline how the analysis can be modified to account for the repeats.

##### 1.3 Proof of convergence

We start by introducing the relevant terminology. Let $\Pi \subset \mathbb{R}^{L+1}$ be a closed set of parameters, and let $f$ be a continuous functional on $\Pi$. Assume that $B \subset \mathbb{R}$ is compact and that $g : \Pi \mapsto \mathcal{C}(B)$ is a continuous mapping from $\Pi$ into $\mathcal{C}(B)$, where $\mathcal{C}(B)$ is the space of continuous functions over $B$ equipped with the supremum norm $||\cdot||_\infty$. For each $D \subset B$ let

$$M(D) = \{\mathbf{c} \in \Pi | \, g(\mathbf{c}, x) \leq 0, x \in D\}$$

denote the set of feasible points of the optimization problem

$$\min f(\mathbf{c}) \text{ over } \mathbf{c} \in M(D).$$

Assuming that $M(D) \neq \emptyset$, let

$$\mu(D) = \inf\{f(\mathbf{c}) | \mathbf{c} \in M(D)\},$$

and define the level set

$$\text{Level}(\mathbf{c}_0, D) = \{\mathbf{c} \in \Pi | \, f(\mathbf{c}) \leq f(\mathbf{c}_0)\} \cap M(D).$$

We also make the following two assumptions:

- Assumption 1: Fine grid Let $\mathbb{N}_0 = \mathbb{N} \cup \{0\}$. There exists a sequence $\{B_i\}$ of compact subsets of $B$ with $B_i \subset B_{i+1}$, $i \in \mathbb{N}_0$, for which $\lim_{i\to\infty} h(B_i, B) = 0$, such that

$$h(B_i, B) = \sup_{x \in B} \inf_{y \in B_i} ||x - y||.$$

- Assumption 2: Bounded level set $M(B)$ is nonempty, and there exists a $\mathbf{c}_0 \in M(B)$ such that the level set $\text{Level}(\mathbf{c}_0, B_0)$ is bounded and hence compact in $\mathbb{R}^{L+1}$.

Convergence of the discretized method, Theorem 2.1 from [1]:

Under assumptions 1.3 and 1.3, the solution of the discretized problem converges to the optimal solution. More formally, we have

$$\mu(B_i) \leq \mu(B_{i+1}) \leq \mu(B), \forall t \in \mathbb{N}_0$$
$$\lim_{i\to\infty} \mu(B_i) = \mu(B).$$

If $\mathbf{c}^*$ is the unique optimal solution of the original problem, and $\mathbf{c}_i^*$ is the optimal solution of the discretized relaxation with grid $B_i$, then

$$\lim_{i\to\infty} ||\mathbf{c}^* - \mathbf{c}_i^*||_2 = 0.$$

It is straightforward to see that our chosen grid is arbitrary fine. Hence, we only need to prove that there exists a $\mathbf{c}_0$ such that the level set $\text{Level}(\mathbf{c}_0, D)$ is bounded.

Let $\mathbf{c} = (\mathbf{a}; t)$ and note that in our setting, $f(\mathbf{c}) = t$. Rewrite $g(\mathbf{c}, \lambda)$ in matrix form as

$$g(\mathbf{c}, \lambda) = \mathbf{a}^T \mathbf{M}(\lambda)\mathbf{a} + \mathbf{a}^T \mathbf{\Lambda}\mathbf{\Lambda}^T \mathbf{a} - t,$$

where

$$\mathbf{\Lambda} \triangleq e^{-\lambda}(\lambda^0, \lambda^1, ..., \lambda^L)^T.$$

Note that only $a_1, ...a_L$ are allowed to vary since we fixed $a_0 = -1$. Obviously, $\mathbf{\Lambda}\mathbf{\Lambda}^T$ is positive semi-definite and the previously introduced $\mathbf{M}(\lambda)$ is positive definite for all $\lambda > 0$. Since the constraints on $g$ are positive definite with respect to $a_1, ...a_L$, $g$ is coercive in $a_1, ...a_L$. Furthermore, for any given $t$, the set of feasible coefficients $a_1, ...a_L$ is bounded. Therefore, given a $t_0$, the level set $\text{Level}(\mathbf{c}_0, B_0)$ is bounded. This ensures that Assumption 1.3 holds for our optimization problem.

Next, we prove the uniqueness of the optimal solution $\mathbf{c}^\star$. Note that proving this result is equivalent to proving the uniqueness of $\mathbf{a}^\star$. Hence, we once again refer to the original minmax formulation of our problem,

$$\inf_{\mathbf{a}:a_0=-1} \sup_{\lambda \in [\frac{n}{k}, 6.5L]} \mathbf{a}^T(\mathbf{M}(\lambda) + \mathbf{\Lambda}\mathbf{\Lambda}^T)\mathbf{a} \triangleq \inf_{\mathbf{a}:a_0=-1} \sup_{\lambda \in [\frac{n}{k}, 6.5L]} h_\lambda(\mathbf{a}) \qquad (2)$$

Clearly, $\forall \lambda \in [\frac{n}{k}, 6.5L]$, the function $h_\lambda(\mathbf{a})$ is strictly convex since $(\mathbf{M}(\lambda) + \mathbf{\Lambda}\mathbf{\Lambda}^T) \succ 0,\ \forall \lambda \in [\frac{n}{k}, 6.5L]$. Taking the supremum over $\lambda$ preserves strict convexity since $\forall \theta \in (0,1)$, one has

$$
\begin{aligned}
&\sup_{\lambda \in [\frac{n}{k}, 6.5L]} h_\lambda(\theta \mathbf{x} + (1-\theta)\mathbf{y}) \\
&< \sup_{\lambda \in [\frac{n}{k}, 6.5L]} \theta h_\lambda(\mathbf{x}) + (1-\theta) h_\lambda(\mathbf{y}) \\
&\leq \sup_{\lambda \in [\frac{n}{k}, 6.5L]} \theta h_\lambda(\mathbf{x}) + \sup_{\lambda' \in [\frac{n}{k}, 6.5L]} (1-\theta) h_{\lambda'}(\mathbf{y}).
\end{aligned}
$$

Hence $\sup_{\lambda \in [\frac{n}{k}, 6.5L]} h_\lambda(\mathbf{a})$ is strictly convex, which consequently implies the uniqueness of $\mathbf{a}^\star$ and hence $\mathbf{c}^\star$.

For the case of samples passed through a Poisson channel, it is not hard to see that the constraints are again strictly convex in $\mathbf{a}$, where one only need to replace $\mathbf{M}(\lambda), \mathbf{\Lambda}$ by

$$
\begin{aligned}
&\frac{1}{k} e^{-\lambda(1-e^{-\eta})}\, Diag(0!\eta^0 M_{N^*}^{(0)}(0), 1!\eta^1 M_{N^*}^{(1)}(0), ..., L!\eta^L M_{N^*}^{(L)}(0)) \\
&e^{-\lambda(1-e^{-\eta})} (\eta^0 M_{N^*}^{(0)}(0), \eta^1 M_{N^*}^{(1)}(0), ..., \eta^L M_{N^*}^{(L)}(0))^T
\end{aligned}
$$

respectively. Thus, a similar analysis is possible and the details are omitted. The proof above along with the previous observation proves the convergence result.

#### 1.4 Proof for the convergence rate

In what follows, and for reasons of simplicity, we omit the constraint $a_0 = -1$ in the SIP formulation. The described proof only requires small modifications to accommodate $a_0 = -1$.

Recall that we used $B_d$ to denote the grid with grid spacing $d$. In order to use the results in [2], we require the convergence assumption below.

- Assumption Let $\bar{\mathbf{c}}$ be a local minimizer of an SIP. There exists a local solution $\mathbf{c}_d$ of the discretized SIP with grid $B_d$ such that

  $$||\mathbf{c}_d - \bar{\mathbf{c}}|| \to 0.$$

  This assumption is satisfied for the SIP of interest as shown in the first part of the proof.

- Assumption The following hold true:
  - There is a neighborhood $\bar{U}$ of $\bar{\mathbf{c}}$ such that the function $\frac{\partial^2}{\partial \lambda^2} g(\mathbf{c}, \lambda)$ is continuous on $\bar{U} \times B$.
  - The set $B$ is compact, nonempty and explicitly given as the solution set of a set of inequalities, $B = \{\lambda \in \mathbb{R} | v_i(\lambda) \leq 0, i \in I\}$, where $I$ is a finite index set and $v_i \in C^2(B)$.
  - For any $\bar{\lambda} \in B$, the vectors $\frac{\partial}{\partial \lambda} v_i(\bar{\lambda}), i \in \{i \in I | v_i(\bar{\lambda}) = 0\}$ are linearly independent.

  Recall that our objective is of the form

  $$g(\mathbf{c}, \lambda) = \mathbf{a}^T \mathbf{M}(\lambda) \mathbf{a} + \mathbf{a}^T \mathbf{\Lambda}\mathbf{\Lambda}^T \mathbf{a} - t,$$

  where

  $$
  \begin{aligned}
  \mathbf{\Lambda} &\triangleq e^{-\lambda}(\lambda^0, \lambda^1, ..., \lambda^L)^T,\ \mathbf{c} = (\mathbf{a}; t), \\
  \mathbf{M}(\lambda) &\triangleq \frac{e^{-\lambda}}{k} Diag(\lambda^0 0!, \lambda^1 1!, ..., \lambda^L L!).
  \end{aligned}
  $$

  It is straightforward to see that the first condition in Assumption 1.4 holds. For the second condition, recall that $B = [\frac{n}{k}, 6.5L]$. Hence, the second condition can be satisfied by choosing $I = \{1\}$, $v_1(\lambda) = (\lambda - \frac{n}{k})(\lambda - 6.5L)$. Since we only have one variable $v_1$, it is also easy to see that the third condition is met.

- Assumption The set $B$ satisfies Assumption 1.4 and all the sets $B_d$ contain the boundary points $\frac{n}{k}, 6.5L$.

  This assumption also clearly holds for the grid of choice. Note that it is crucial to include the boundary points for the proof in [2] to be applicable.

- Assumption $\nabla_{\mathbf{c}} g(\mathbf{c}, \lambda)$ is continuous on $\bar{U} \times B$, where $\bar{U}$ is a neighborhood of $\bar{\mathbf{c}}$. Moreover, there exists a vector $\xi$ such that

  $$\nabla_{\mathbf{c}} g(\bar{\mathbf{c}}, \lambda)^T \xi \leq -1, \ \forall \lambda \in B.$$

  Note that $\nabla_{\mathbf{c}} g(\mathbf{c}, \lambda) = [\nabla_{\mathbf{a}} g(\mathbf{c}, \lambda); \nabla_t g(\mathbf{c}, \lambda)]$ and

  $$\nabla_{\mathbf{a}} g(\mathbf{c}, \lambda) = 2(\mathbf{M}(\lambda) + \mathbf{\Lambda}\mathbf{\Lambda}^T)\mathbf{a}.$$

  Also note that $\forall \lambda \in B$, $\mathbf{M}(\lambda) + \mathbf{\Lambda}\mathbf{\Lambda}^T$ is positive definite. Hence choosing $\xi$ to be colinear with and of the same direction as $[-\mathbf{a}^T \ 1]^T$, as well as of sufficiently large norm will allow us to satisfy the inequality

  $$\nabla_{\mathbf{c}} g(\bar{\mathbf{c}}, \lambda)^T \xi \leq -1, \ \forall \lambda \in B.$$

  Hence, Assumption 1.4 holds as well. The next results follow from the above assumptions and observations, and the results in [2].

Corollary 1 in [2]
Let $t_d$ be the optimal objective value of the discretized SIP used for support estimation with the grid $B_d$, and let $t^\star$ be the optimal objective value for the original SIP. Since Assumptions 1.4,1.4,1.4,1.4 hold, then for some $c_3 > 0$ and $d$ sufficiently small, we have

$$0 \leq t^\star - t_d \leq c_3 d^2.$$

Consequently, $t_d \to t^\star$ with a convergence rate of $O(d^2)$.
Theorem 2 in [2]
Assume that all assumptions in Lemma 1.4 hold. If there exists a constant $c_4 > 0$ such that

$$t - \bar{t} \geq c_4 ||\mathbf{c} - \bar{\mathbf{c}}||, \ \forall \mathbf{c} \in M(B) \cap \bar{U},$$

then for sufficiently small $d$ and $\sigma > 0$ we have

$$||\mathbf{c}_d - \bar{\mathbf{c}}|| \leq \sigma d^2.$$

This result implies that if $\bar{\mathbf{c}}$ is also a strict minimum of order one, then the solution of the discretized SIP converges to that of the the original SIP with rate $O(d^2)$. For the Poisson repeat channel, the constraints are also strictly convex in $\mathbf{a}$. Therefore, a similar analysis is possible and the details are omitted once again. Combining these results completes the proof.

#### 1.5 Additional theoretical results

The result described in the main text follows from Theorem 6.2 in [3].

Theorem 6.2 from [3] Let $W(x) = \exp(-Q(x))$ be a weight function, where $Q : \mathbb{R} \mapsto [0, \infty)$ is even, convex, diverging for $x \to \infty$, and such that

$$0 = Q(0) < Q(x), \forall x \neq 0.$$

Then, for any polynomial $P(x)$ of degree $\leq L$, not identical to zero, one has

$$\sup_{x \in \mathbb{R}} |P(x)W(x)| = \sup_{x \in [-M_L, M_L]} |P(x)W(x)|,$$
$$\sup_{x \in \mathbb{R} \backslash [-M_L, M_L]} |P(x)W(x)| < \sup_{x \in [-M_L, M_L]} |P(x)W(x)|.$$

Here, $M_L$ stands for the *Mhaskar-Rakhmanov-Saff* (MSF) number, which is the smallest positive root of the integral equation

$$L = \frac{2}{\pi} \int_0^1 \frac{M_L t Q'(M_L t)}{\sqrt{1-t^2}} dt. \tag{3}$$

In our setting, the weight equals $\exp(-x)$. Solving (3) gives us an MSF number equal to $M_L = \frac{\pi}{2} L$. Thus, we can restrict our optimization interval to $[\frac{n}{k}, \frac{\pi}{2} L + \frac{n}{k}]$. If there is no regularization term, the optimal interval reduces to $[\frac{n}{k}, \frac{\pi}{2} L + \frac{n}{k}]$.

##### 1.6 Construction of the RWC-S estimator

We introduce the optimization problem needed for minimizing the risk $E\left(\frac{S-\hat{S}}{S}\right)^2$. Poissonization arguments once again establish that

$$\mathbb{E}\left(\frac{S-\hat{S}}{S}\right)^2 = \frac{1}{S^2}\Big\{\sum_{i\in\mathcal{L}}\Big(\sum_{l=0}^{L} e^{-\lambda_i} a_l^2 \lambda_i^l l!\Big) + \sum_{i\neq j\in\mathcal{L}}\Big(e^{-\lambda_i}\sum_{l=0}^{L} a_l\lambda_i^l\Big)\Big(e^{-\lambda_j}\sum_{l=0}^{L} a_l\lambda_j^l\Big)\Big\}.$$

Taking the supremum over $D_k$, one can further upper bound the risk as

$$\begin{aligned}
&\leq \sup_{\lambda_\ell\in[\frac{n}{k},n],\ \ell\in\mathcal{L}} \frac{1}{S^2}\Big\{\sum_{i\in\mathcal{L}}\Big(\sum_{l=0}^{L} e^{-\lambda_i} a_l^2 \lambda_i^l l!\Big) + \sum_{i\neq j\in\mathcal{L}}\Big(e^{-\lambda_i}\sum_{l=0}^{L} a_l\lambda_i^l\Big)\Big(e^{-\lambda_j}\sum_{l=0}^{L} a_l\lambda_j^l\Big)\Big\} \\
&\leq \sup_{\lambda\in[\frac{n}{k},n]} \left\{\frac{1}{S}\Big(\sum_{l=0}^{L} e^{-\lambda} a_l^2\lambda^l l!\Big) + \Big(e^{-\lambda}\sum_{l=0}^{L} a_l\lambda^l\Big)^2\right\} \\
&\leq \sup_{\lambda\in[\frac{n}{k},n]} \left\{\frac{1}{\hat{S}_c}\Big(\sum_{l=0}^{L} e^{-\lambda} a_l^2\lambda^l l!\Big) + \Big(e^{-\lambda}\sum_{l=0}^{L} a_l\lambda^l\Big)^2\right\}, \qquad (4)
\end{aligned}$$

where the last inequality is due to the fact that $\hat{S}_c \leq S$. Note that the only difference between (4) and the corresponding optimization problem described in the main text is in terms of changing the normalization from $1/k$ to $1/\hat{S}_c$ in the first term. The expression (4) is optimized by the solution of the following problem:

$$\begin{aligned}
&\min_{t,\mathbf{a}\in Poly(L)} \quad t \quad \text{s.t.} \\
&\left\{\frac{1}{\hat{S}_c}\Big(\sum_{l=0}^{L} e^{-\lambda} a_l^2\lambda^l l!\Big) + \Big(e^{-\lambda}\sum_{l=0}^{L} a_l\lambda^l\Big)^2\right\} \leq t,\ \forall\lambda\in \text{Grid}([\frac{n}{k}, 6.5L], s).
\end{aligned} \qquad (5)$$

### 2 Performance on non-iid data with ground truth

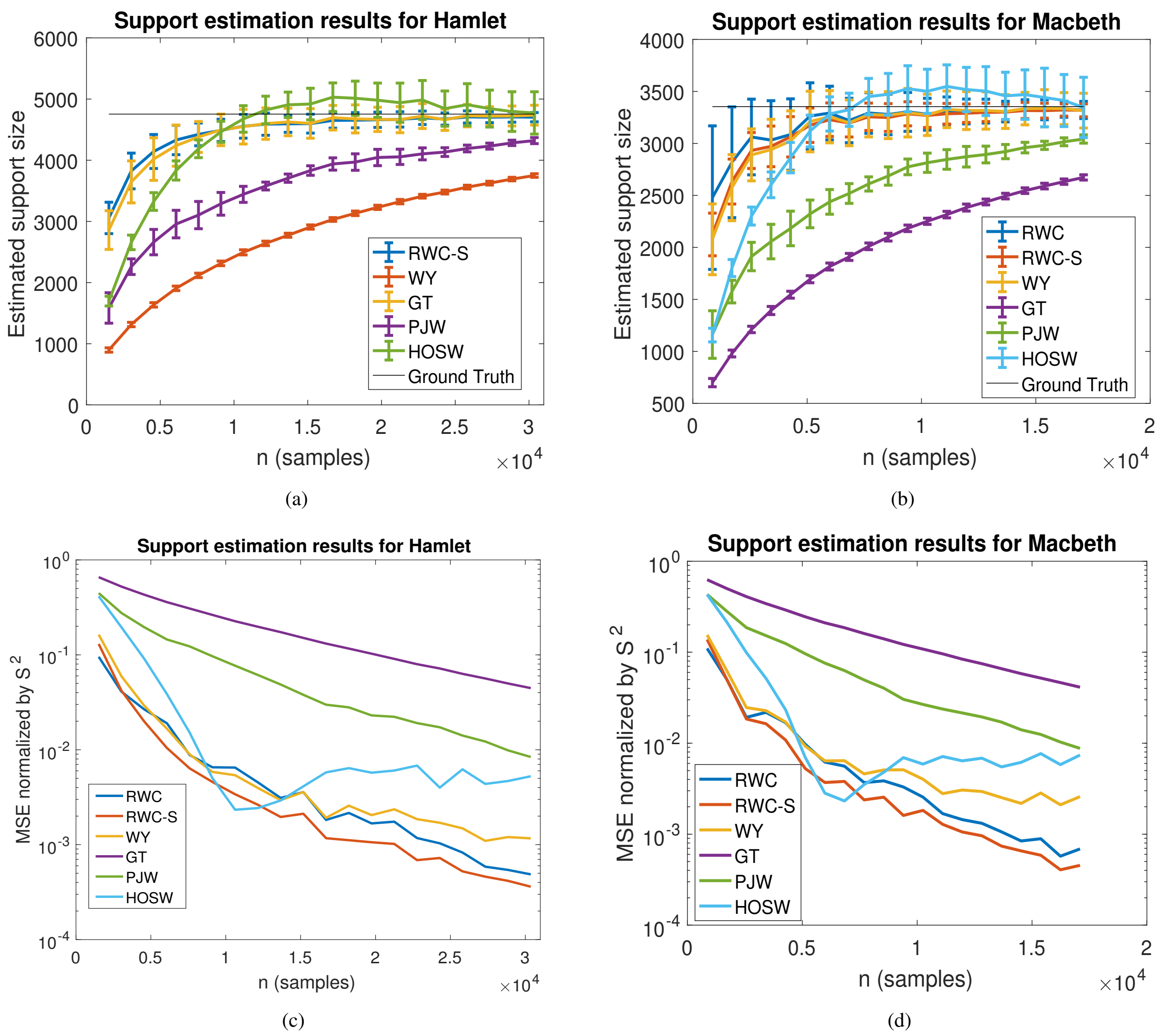


Figure S1: The results are obtained over $100$ independent trials. The first two figures shows the mean and standard deviation of the estimators, while the latter two figures shows the MSE normalized by $S^2$.
